# Individualized cortical thickness asymmetry in Autism Spectrum Disorders and Schizophrenia

**DOI:** 10.1101/2024.11.06.24316751

**Authors:** Marta Martin Echave, Hugo G. Schnack, Covadonga M. Díaz-Caneja, Laura Pina-Camacho, Niels Janssen, Pedro M. Gordaliza, Kuan H. Kho, Elizabeth E.L. Buimer, Neeltje E.M. van Haren, René S. Kahn, Hilleke E. Hulshoff Pol, Mara Parellada, Celso Arango, Joost Janssen

## Abstract

**Introduction:** Cortical thickness asymmetry has been proposed as a latent biomarker for Autism Spectrum Disorders (ASD) and schizophrenia (SZ). However, the degree of abnormal asymmetry at the individual level in ASD and SZ remains unclear. To investigate this, we applied normative modeling.

**Methods:** Normative means for the whole brain and regional (160 cortical parcels) cortical thickness asymmetry index (AI) were established using a training set of healthy subjects (n=4,904, 45.15% male, age range: 6-95 years), controlling for age, sex, image quality and scanner. We calculated z-scores to quantify individual deviations from the normative mean in a test set consisting of healthy controls (HC_test_, n=526, 40% male), participants with ASD (n=135, 83% male) and SZ (n=287, 81% male). Regional deviance was assessed by counting the number of individuals with significant deviations below (infra-normal, z-score ≤ -1.96) or above (supra-normal, z-score ≥ 1.96) normative means in each parcel. We also evaluated individual deviance by counting the number of regions with significant deviations for each participant. A data-driven multivariate approach was employed to determine whether joint regional deviance was associated with diagnosis.

**Results:** There were no differences for deviance of whole brain AI between any of the groups. Distributions of individual deviances overlapped across all 160 regions, with only one superior temporal region in which SZ individuals showed a higher proportion of supra-normal AI values compared to HC_test_ (HC_test_ = 1.14%, SZ = 5.92%, **χ**2 = 15.45, P_FDR_< 0.05, ω = 0.14). The SZ group also had a higher average number of regions with significant deviations than HC_test_ (infra-normal: z = -4.21, p < 0.01; supra-normal: z = -4.33, p < 0.01). Multivariate analysis showed no association between inter-regional heterogeneity of AI and diagnosis. Results were consistent when using a higher resolution parcellation, alternative asymmetry calculations, analysis restricted to males, and after controlling for handedness and IQ.

**Conclusions:** Our findings indicate that whole brain, regional and inter-regional variability in cortical thickness AI among those with ASD is entirely accounted for by normative variation. This study challenges the utility of cortical thickness asymmetry as a biomarker for ASD.

## Introduction

The cortical thickness of the human cortex exhibits hemispheric asymmetry in its structural organization (Zhou et al., 2013). Case-control studies in schizophrenia (SZ) and autism spectrum disorders (ASD) indicate that individuals with these conditions often lack the typical leftward asymmetry in language-related brain regions, where the left hemisphere is usually larger than the right (Dougherty et al., 2016; Floris et al., 2016). Notably, SZ and ASD are also associated with pronounced language deficits and possibly increased rates of non-right-handedness (Hirnstein and Hugdahl, 2014). Collectively, these findings suggest that hemispheric asymmetry of brain structure may play a role in the neurobiology of ASD and SZ and hold potential as a biomarker for these conditions (Ratnanather et al., 2013; Li et al., 2023).

However, recent large-scale case-control imaging studies from the ENIGMA initiative present conflicting results regarding the significance and extent of hemispheric asymmetry in ASD and SZ (Kong et al., 2022). These studies reveal a high degree of inter-individual variability of brain asymmetry in both ASD and SZ, challenging the notion of consistent hemispheric asymmetry across individuals with ASD or SZ (Kong et al., 2022). To specifically address individual phenotypic heterogeneity, normative modeling frameworks have been developed, which establish standard norms for neurobiological variables and assess individual deviations from these benchmarks (Marquand et al., 2019). Applying normative modeling to cortical thickness in ASD and SZ has revealed that individual deviations in regional cortical thickness do not occur consistently in the same regions or with the same severity (Zabihi et al., 2019; Bethlehem et al., 2020; Lv et al., 2021; Di Biase et al., 2022; Segal et al., 2023). However, no studies have yet applied this analytical approach to regional cortical thickness asymmetry, leaving it unclear whether deviations from normative cortical thickness asymmetry are disproportionately represented among individuals with ASD or SZ. This study’s aims to assess and analyze the heterogeneity of regional cortical thickness asymmetry in a large, multi-scanner sample of healthy individuals as well as those with ASD or SZ.

## Methods

### Sample characteristics

We examined data for 5430 healthy individuals (44.68% male) and 422 cases, taken from 20 different scan sites. The clinical sample comprised 135 individuals with ASD (83% male) and 287 individuals with SZ (81% male). The age distributions per scan site are given in Supplemental Figure 1). The scanner details, sample size, demographic and clinical characteristics of each scan site, after various exclusions based on data quality are presented in Supplemental Table 1.

### Image processing and quality control

For a flowchart leading to the final sample, see Supplemental Figure 2. The ABIDE-I and ABIDE-II datasets include low quality T1 data which biases cortical thickness measurements (Bedford et al., 2023). We initially included only images from ABIDE that we denoted as the highest quality (Bedford et al., 2023). The BGS and COBRE datasets have undergone extensive quality control procedures (Chopra et al., 2023); we initially included the same images from these two samples as (Chopra et al., 2023). For the Utrecht dataset images also underwent extensive quality control and we initially included the same images as (Janssen et al., 2021). Thereafter, using the initially included raw T1-weighted images from all datasets we applied the Computational Anatomy Toolbox to generate a weighted overall image quality rating (IQR) for every image (Gaser et al., 2024). This metric combines ratings of basic image properties, including the level of noise and geometric distortions, into a single score that quantifies the overall image quality of a participant’s T1-weighted scan (for more information, see Gaser et al. 2024). On this metric, lower scores denote higher image quality. As per previous work, we excluded 682 images with an IQR >2.8 (Wolfers et al., 2018). Thereafter all remaining images were processed centrally using the FreeSurfer analysis suite (v7.1) with default settings (Fischl, 2012). The Freesurfer Euler number was extracted as a proxy for freesurfer surface quality (Rosen et al., 2018). Finally, as per previous work, images were removed if the maximum, absolute, within-dataset centered Euler number was larger than 10 (Rutherford et al., 2023). This was the case for five images.

### Cortical thickness hemispheric Asymmetry Index

#### Whole brain

We used the average cortical thickness for the left (LH) and right hemisphere (RH) as outputted by FreeSurfer’s reconstruction pipeline. We then computed the whole brain hemispheric Asymmetry Index (AI) for cortical thickness for each subject (n) as follows:

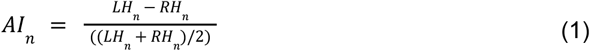

where *LH*_*n*_ represents the average cortical thickness for subject *n* for the left hemisphere and *RH*_*n*_ represents the average cortical thickness for subject *n* for the right hemisphere. A positive AI value reflects leftward asymmetry (LH > RH).

#### Region

We applied a validated symmetric parcellation of 160 regions of approximately equal size to the individual cortical thickness maps from the left and right hemisphere outputted by FreeSurfer’s reconstruction pipeline (Romero-Garcia et al., 2014). For each left and right hemispheric region we calculated the cortical thickness. We then computed the AI for cortical thickness for each hemispheric region (r) and each subject (n) as follows:

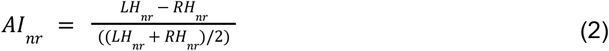

where *LH*_*nr*_ represents the cortical thickness for subject *n* and region *r* for the left hemisphere and *RH*_*nr*_ represents the cortical thickness for subject *n* and corresponding region *r* for the right hemisphere.

### Alternative approaches

#### Alternative A1: Using a higher resolution atlas

Given that other studies calculated AI at higher spatial resolution, we processed all images using a validated higher resolution parcellation (1000 regions) and calculated the AI for each region (Schaefer et al., 2018; Kruggel and Solodkin, 2020; Roe et al., 2023).

#### Alternative A2: Using alternative image processing for calculating AI

We generated unsmoothed vertex-wise standard space cortical thickness maps for each participant and hemisphere from Freesurfer using Freesurfer’s ‘qcache’ command. These maps were inserted into Freesurfer’s ‘xhemi’, a dedicated image processing pipeline for cortical asymmetry analysis (Greve et al., 2013). The procedure achieves alignment of the homotopic hemisphere by cross-hemispheric registration using a symmetric surface template (Roe et al., 2023). In our case, we mapped the right hemisphere to the left hemisphere. We then computed the AI for cortical thickness at each left hemispheric vertex (v) and each subject (n) as follows:

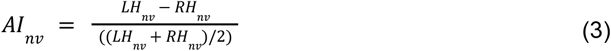

where *LH*_*nv*_ represents the cortical thickness for subject *n* and vertex *v* for the left hemisphere and *RH*_*nv*_ represents the cortical thickness for subject *n* and vertex *v* for the right hemisphere. To reduce the computational burden for subsequent normative modeling and maximize comparability with our main approach we parcellated the left hemisphere of each participant into the same 160 similar sized regions as in our main approach. The AI was subsequently calculated by averaging across vertices belonging to a region.

#### Alternative A3: Using males only

We calculated the whole brain and regional AI using males only. Separate analyses in females were not possible due to insufficient sample size.

### Normative modeling

For each of the datasets exclusively comprising healthy controls 90% of individuals went into the training set and 10% into the test set (HC_test_) using the createDataPartition function from the caret R package. We maintained the distribution of age, sex, and scanner variables between the training and test sets. From each of the clinical datasets, i.e. those datasets including cases and healthy controls, 90% of the healthy controls were added to the training set and 10% were added to the HC_test_. All cases with ASD or SZ were added to the test set. Both the training and test sets comprised individuals from the same scanners, constituting a “within-site-split”.

Subsequently, we used Bayesian Linear Regression (Fraza et al., 2021). The training set consisted of 4904 healthy individuals (54.85% female, age range: 6-95 years) which established a normative range for AI, considering an individual’s age, sex, Euler number, and scanner. The age range encompassed the age range of the cases (range: 7-65 years). For each region, we assessed the extent to which the AI estimate, for each individual, deviated from the predictions of the normative model. These deviations were quantified as z-scores, for each region (r) and each subject (n), as follows:

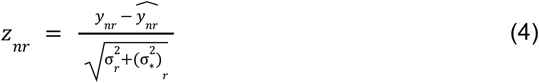

where *y*_*nr*_ means the true AI and *ŷ*_*rs*_ is the predicted average AI. The difference in these values is normalized to account for two different sources of variation; i) 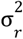 which is the aleatoric uncertainty and reflects the variation between individuals across the population, and ii) he epistemic uncertainty, 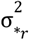, which accounts for the variance associated to modeling uncertainty introduced by the model structure or parameter selection (associated to the age gaps in which there is low density of individuals). As per prior publications we considered AI z-scores as infra- or supra-normal when they equalled or exceeded -1.96 or 1.96, respectively (Bethlehem et al., 2020; Lv et al., 2021; Di Biase et al., 2022; Rutherford et al., 2023; ENIGMA Clinical High Risk for Psychosis Working Group et al., 2024; Huang et al., 2024).

To evaluate the generalization of our model, we employed a ten-fold cross-validation approach on the training cohort. Specifically, we divided the training cohort into ten separate folds. In each fold, we trained BLR models using 90% of the participants as training data, while including age, sex, Euler number, and scanner as covariates, reserving the remaining 10% for assessing generalization performance. This process was repeated ten times so that predictions of AI values were generated for all individuals within the training cohort group. This methodology follows standard practices in machine learning and yields nearly unbiased estimates of the model’s true generalization capabilities. Z-scores were calculated and we assessed model fit by calculating the root-mean squared error between predicted and true AI values for our main approach and the three alternative approaches (A1, A2 and A3), see Supplemental Figures 3, 5 and 10.

We assessed data bias over sites using linear support vector classifiers. Specifically, we employed a series of one-versus-all linear support vector machines, each with a default slack parameter of 1 (Segal et al., 2023). These models were trained separately on the z-scores from the healthy controls from the training subsets and the test subsets for the purpose of classifying scan sites. For each site, a two-fold linear support vector machine classifier was trained and the mean balanced accuracy was calculated. A mean balanced accuracy near chance level (50%) served as an indicator that the observed deviations were minimally influenced by residual site effects, which was confirmed for our main approach and the A1 and A2 approaches (see Supplemental Tables 2-4).

### Statistical analyses

#### Normative modeling-based deviance per region

For each region we calculated, separately for HC_test_ and each disorder, the percentages of individuals with infra- and supra-normal outlier values, see Figure 1: A-D. This led to the creation of percentage outlier maps to visualize the spatial distribution, see Figure 1: E. To mitigate the impact of varying sample sizes among groups, we utilized the percentages of individuals rather than raw counts. Group differences in the proportion of individuals with supra- or infra-normal z-scores between HC_test_ and each disorder were examined using permuted chi-square tests (n=10000) using the coin R package with FDR (q = 0.05) as a correction for multiple comparisons and Cohen’s ω (which has an identical interpretation as Cohen’s d) as a measure of effect size.

**Figure 1.**
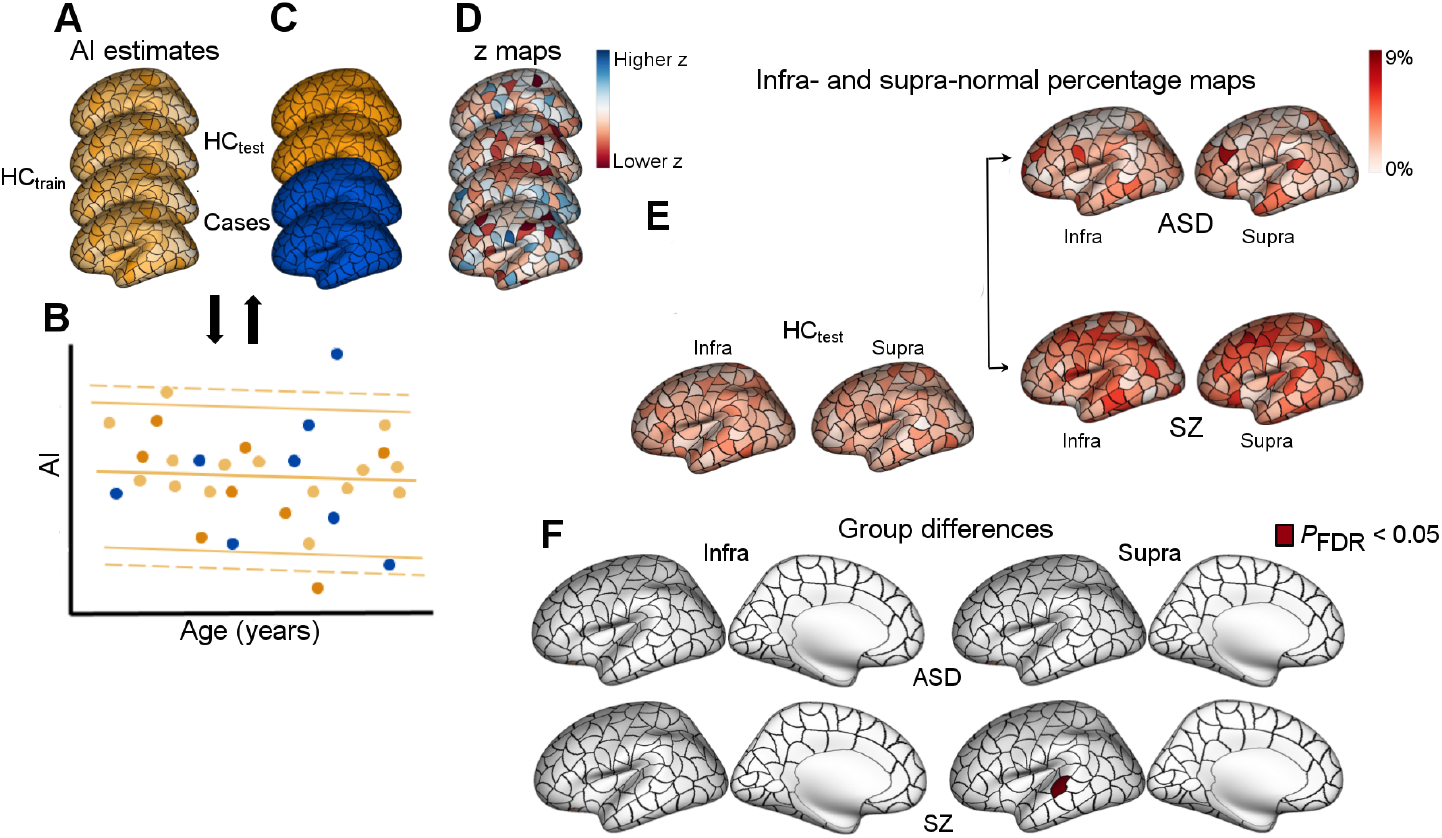
Characterizing regional-level deviance in the Asymmetry Index (AI). The cortex for each individual was parcellated into 160 equally sized cortical regions (Romero-Garcia et al., 2014) (**A**). The training dataset, HC_train_, was used to train a normative model to make predictions about regional AI values given an individual’s age, sex, Euler number, and scan site (**B**). The predictions for held-out healthy individuals (HC_test_) and cases were then compared with empirical AI estimates. Model predictions for one region, showing individuals in the training set (HC_train_; light orange) and the held-out controls (HC_test_; dark orange) and clinical groups (blue). (**C**). For each individual, deviations from model predictions were quantified as a z-score map (**D**). For the HC_test_ and each clinical group, we quantified the proportion of individuals showing an infra- and supra-normal deviation in a given brain region, yielding infra- and supra-normal deviation percentage maps (**E**). We compared the percentage maps from HC_test_ and each clinical group using chi-square permutation tests and FDR correction yielding a map showing regions with significantly different percentages of infra- and supra-normal AI deviations in ASD or SZ compared with HC_test_ (**F**). AI, asymmetry index; HC_train_, healthy control individuals from the training set; HC_test_, healthy control individuals from the test set; ASD, Autism Spectrum Disorders; SZ, schizophrenia. Data used to generate this figure can be found in the Supplemental Data.

#### Normative modeling-based deviance per individual

##### Whole brain

The whole brain cortical thickness AI from each individual in the test set was used to calculate their z-score, representing the individual deviance for whole brain cortical thickness AI per individual. Group differences in z-scores for whole brain cortical thickness AI between HC_test_ and each disorder were examined using Welch t tests and Cohen’s d was used as a measure of effect size. Group differences in the proportion of individuals with supra- or infra-normal z-scores between HC_test_ and each disorder were examined using permuted chi-square tests (n=10000) using the coin R package and Cohen’s ω was used as a measure of effect size.

##### Region

The regional cortical thickness AI from each individual in the test set was used to calculate their z-score, representing the individual deviance for regional cortical thickness AI. For each individual we calculated the number of regions with infra- and supra-normal outlier values. Group differences in the number of regions with supra- or infra-normal z-scores between HC_test_ and each disorder were examined using the Mann-Whitney U test.

#### Multivariate analysis of normative modeling-based deviance

We finally investigated whether the multivariate pattern of regional cortical thickness AI, represented by the z-scores of all regions for each participant, revealed any diagnostic effect. To explore this, we conducted an exploratory clustering analysis to determine if the z-scores across the entire cortex could distinguish between HC_test_, ASD, and SZ groups. A data-driven clustering approach was applied, using t-Distributed Stochastic Neighbour Embedding (tSNE) to construct a distance matrix from the 160 regions by 948 participants’ (all participants included in the test set) z-scores. K-medoid clustering was then performed on this distance matrix, grouping participants into maximally independent clusters based on the optimal number of clusters determined by average silhouette width (Hennig and Liao, 2013). Finally, k-medoid clustering was re-run with the optimal cluster count, and the overlap between clusters and diagnoses was visualized by projecting the clustering onto the 2-dimensional tSNE-embedded space.

#### Replication analyses

To assess the robustness of our results we repeated all analyses using AI data derived from the higher resolution parcellation (A1), using the AI derived from the alternative image processing pipeline (A2) and using males only (A3). Fourthly, we assessed the effect of handedness and IQ on the average z-scores of each individual within HC_test_ and each disorder. Handedness information was available for 238 individuals with SZ and 69 individuals with ASD. Group differences between left- and right-handers in average z-scores were examined using Welch t tests within HC_test_ and each disorder group. Intelligence Quotient (IQ) information was available for 110 individuals with SZ and 69 individuals with ASD. The relationship between IQ and average z-scores was assessed using Pearson correlation within each disorder group.

## Results

### Normative modeling-based deviance per region

Here we counted for each region the number of individuals who had infra- or supra-normal z-scores. The percentages of ASD, SZ and HC_test_ with infra- and supra-normal z-scores for each region and whether the differences between HC_test_ and ASD and HC_test_ and SZ were significant are included in the Supplemental Data. For all the regions the distributions of the z-scores of the ASD and SZ groups overlapped with HC_test_, see Supplemental Figure 4. There were no significant group differences in the proportion of individuals with infra-normal or supra-normal z-scores between HC_test_ and ASD. There were no significant differences in the proportion of individuals with infra-normal z-scores between HC_test_ and SZ. Out of 160 regions, there was one region in the superior temporal cortex, superior temporal cortex part 1, where the proportion of individuals with supra-normal z-scores differed significantly between HC_test_ and SZ (HC_test_ = 1.14%, SZ = 5.92%, **χ**^2^ = 15.45, P_FDR_< 0.05, ω = 0.14), see Figure 1: F. When assessing the mean AI in the superior temporal cortex part 1, the groups of individuals from HC_test_ and SZ with supra-normal z-scores in the superior temporal cortex part 1 both showed leftward asymmetry and the two groups did not differ from each other in mean ‘raw’ AI (mean AI individuals from HC_test_ with supra-normal z-scores in the superior temporal cortex part 1 = 0.15, mean AI individuals from SZ with supra-normal z-scores in the superior temporal cortex part 1 = 0.16, t= 0.11, P > 0.05, d = 0.07).

The minimum and maximum proportion of infra-normal deviance for any region for individuals with ASD was 0% to 8.15%; 0.7% to 6.7% for SZ; and 1.33% to 4.75% for HC_test_; the corresponding ranges for the minimum and maximum proportion of supra-normal deviance for any region was 0% to 8.89% for ASD; 0.7% to 7.31% for SZ; and 0.76% to 4.56% for HC_test_.

### Normative modeling-based deviance per individual

#### Whole brain

Here we calculated the z-scores for the whole brain AI and counted the number of individuals who had infra- or supra-normal deviance. The distributions of the z-scores of the whole brain AI overlapped for HC_test_, ASD, and SZ groups, see Figure 2. There were no differences in z-scores for whole brain AI between HC_test_ and ASD (P > 0.05, Cohen’s d (d) = -0.01) and between HC_test_ and SZ (P > 0.05, d = -0.02). There were no differences in the proportion of individuals with infra-normal or supra-normal deviance between HC_test_ and ASD for whole brain AI (proportion of individuals with infra-normal z-scores: HC_test_ = 2.47%, ASD = 1.48%, P > 0.05, Cohen’s ω (ω) = 0.05; proportion of individuals with supra-normal z-scores: HC_test_ = 2.66%, ASD = 5.93%, P > 0.05, ω = 0.15) and between HC_test_ and SZ (proportion of individuals with infra-normal z-scores: SZ = 3.48%, P > 0.05, ω = 0.06; proportion of individuals with supra-normal z-scores: SZ = 3.14%, P > 0.05, ω = 0.03).

**Figure 2.**
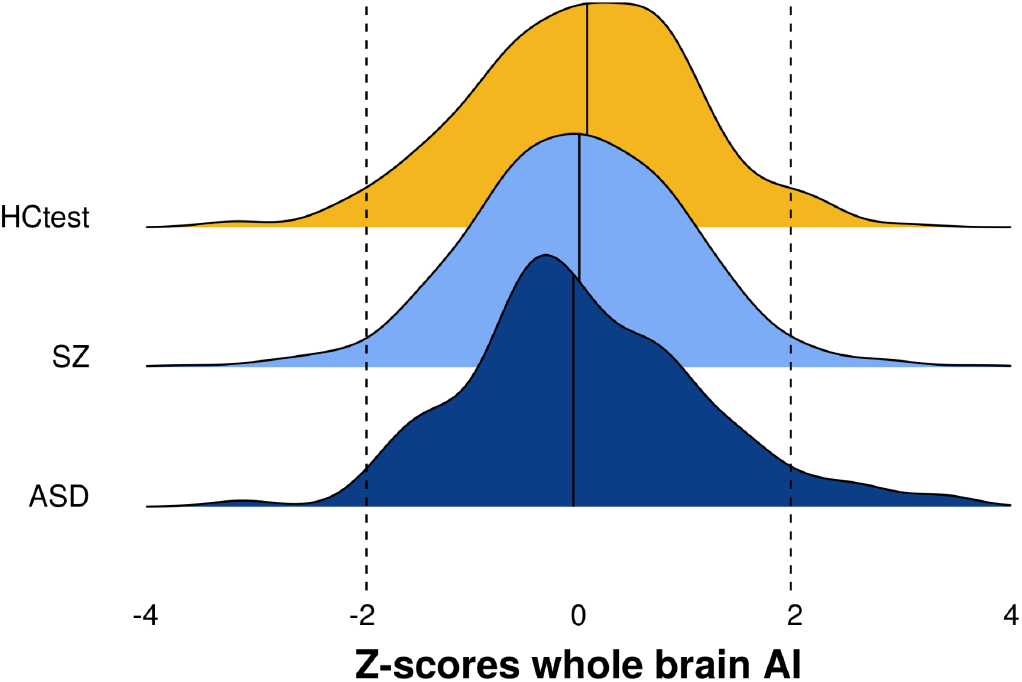
The distributions of the normative modeling based z-scores for the whole brain AI in HC_test_, SZ, and ASD. The vertical line represents the median for each group. The dotted lines represent |z| = 1.96. AI, Asymmetry Index; HC_test_, healthy controls from the test set; ASD, Autism Spectrum Disorders; SZ, schizophrenia.

#### Region

Here we calculated the z-scores for the AI of each region and counted for each individual the number of regions with infra- or supra-normal deviance. The distributions of the proportion of individuals with regional infra- and supra-normal deviance were similar for ASD and HC_test_; there were no group differences in the average number of infra- and supra-normal regions between ASD and HC_test_, see Figure 3. The SZ group had a higher average number of infra- and supra-normal regions compared to HC_test_ (infra-normal: z=-4.21, p<0.01, supra-normal: z=-4.33, p<0.01), see Figure 3. Infra-normal z-scores for AI in at least one region were observed in 93%, 96% and 92% of individuals across ASD, SZ, and HC_test_, respectively; the corresponding supra-normal z-scores were 93%, 97% and 96% across ASD, SZ, HC_test_, respectively.

**Figure 3.**
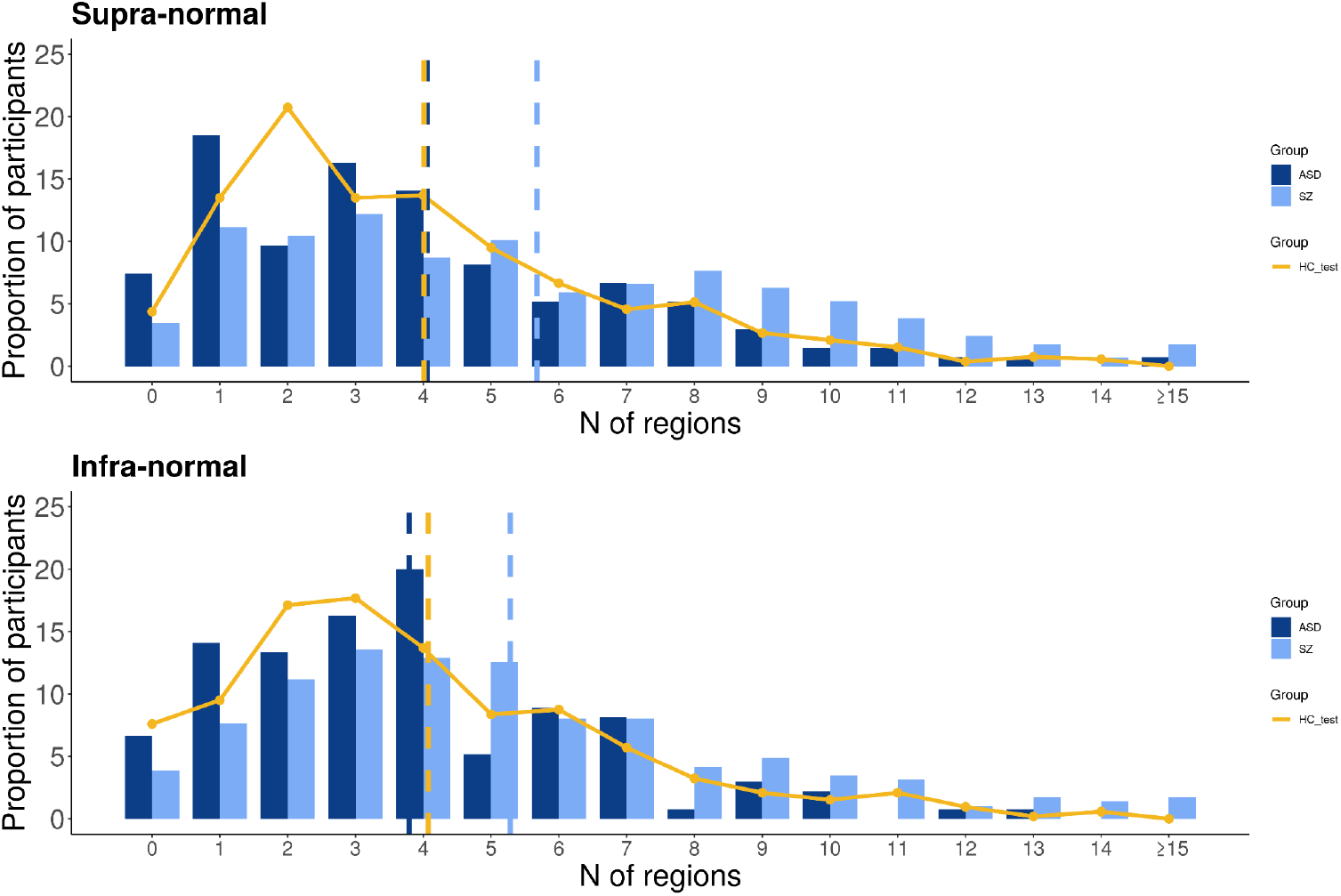
Distribution of the number of regions with infra- or supra-normal deviance per individual. Bars and curves display the distribution of the proportion of individuals per amount of regions with supra-normal and infra-normal deviations. For example, approximately 7% of the individuals with ASD is not supra-normal deviant in any region while approximately 15% of the individuals with ASD is supra-normal deviant in one region only. Vertical dashed lines display the average number of infra- and supra-normal regions for each group. HC_test_, healthy individuals from the test set; ASD, Autism Spectrum Disorders; SZ, schizophrenia.

#### Multivariate analysis of normative modeling-based deviance

The results of clustering the neighbor-embedded z-scores for regional AI are displayed in Figure 4. As shown in panel B, there is no significant relationship between the clustering of z-scores and diagnosis. In other words, the within-group heterogeneity is reflected by normative heterogeneity, and the overall density plots for the three groups appear nearly identical.

**Figure 4.**
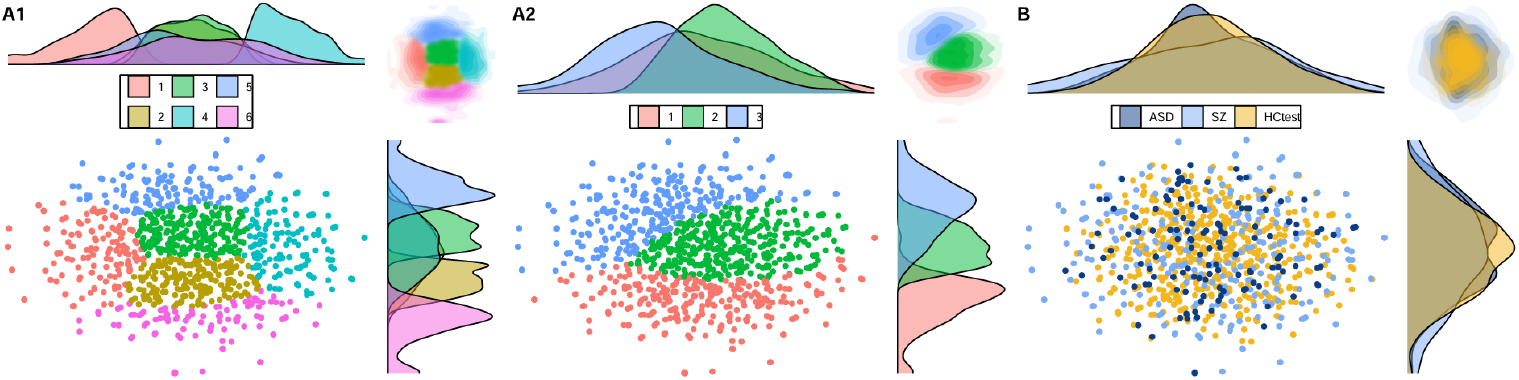
Panels A1 and A2 present the optimum (A1) and three cluster results (A2) of k-medoid clustering applied to the 2D embedding of z-scores for regional cortical thickness AI, generated using tSNE. Panel B demonstrates that these clusters did not yield any meaningful differentiation based on diagnosis. AI, Asymmetry Index; HC_test_, healthy individuals from the test set; ASD, Autism Spectrum Disorders: SZ, schizophrenia.

### Replication analyses

Approaches using a higher resolution parcellation (A1), using an alternative processing method for calculating the AI (A2), and assessing only males (A3) confirmed the results from the main approach.

#### Normative modeling-based deviance per region

For A1, A2 and A3 the distributions of the regional z-scores of the ASD, SZ groups overlapped with HC_test_, see Supplemental Figures 6, 11 and 16. For A1, A2, and A3 there were no significant group differences in the proportion of individuals with regional infra-normal or supra-normal z-scores between HC_test_ and ASD, see Supplemental Data, Supplemental Text and Supplemental Figures 7, 12, and 17. For A1, in less than 2% of the total number of regions, the SZ group had an increased percentage of individuals with infra-normal deviance, see Supplemental Data, Supplemental Text and Supplemental Figure 7. For A2, there was one region located in the medial orbitofrontal cortex where the SZ group had an increased percentage of individuals with infra-normal deviance, see Supplemental Data, Supplemental Text and Supplemental Figure 12. For A3 there were no significant group differences in the proportion of individuals with infra-normal or supra-normal z-scores between HC_test_ and SZ, see Supplemental Data and Supplemental Text and Supplemental Figure 17.

#### Normative modeling-based deviance per individual

##### Whole brain

Whole brain cortical thickness AI for A1 is identical to the main approach. A2 and A3 showed no significant group differences for deviance of whole brain cortical thickness AI, see Supplemental Text and Supplemental Figures 13 and 18.

##### Region

For A1, A2, and A3 there were no significant group differences in the number of infra- and supra-normal regions between HC_test_ and ASD. For A1, A2, and A3 the SZ group had a higher mean number of infra- and supra-normal regions compared to HC_test_, see Supplemental Text and Supplemental Figures 8, 14, and 19.

#### Multivariate analysis of normative modeling-based deviance

For A1, A2 and A3 there were significant relationships between the clustering of z-scores and diagnosis, see Supplemental Figures 9, 15 and 20.

#### Handedness and IQ

Average z-scores of left- and right-handers did not differ within HC_test_ and each disorder group (P > 0.05). There was no significant correlation between average z-scores and IQ in both disorder groups (P > 0.05).

## Discussion

The main finding of this study is that variation in regional asymmetry of cortical thickness in individuals with ASD falls within the normal inter-individual variation. Whole brain and regional supra- and infra-normal deviations of asymmetry of cortical thickness are present in similar proportions in individuals with ASD, SZ and healthy individuals. For individuals with SZ there was one region in the superior temporal cortex where individuals with SZ had a higher proportion of supra-normal z-scores compared to healthy individuals, with very low effect sizes. Prior studies have shown regional cortical thickness abnormalities in SZ and ASD when compared to healthy individuals (van Erp et al., 2018; Pretzsch and Ecker, 2023; Bedford et al., 2024). Our study shows that individual deficits for whole brain and regional cortical thickness asymmetry, defined through normative modeling, are generally absent in both disorders.

Recent case-control studies using large sample sizes from the ENIGMA consortium assessing regional cortical thickness asymmetry in ASD, SZ, and healthy individuals have shown that significant case-control differences of AI were subtle and not widespread, affecting 9% of regions in ASD and 3% of regions in SZ (Postema et al., 2019; Kong et al., 2022; Schijven et al., 2023). Floris et al. (2021) is the only study, to the best of our knowledge, to use the normative modeling framework for assessing structural brain asymmetry in ASD and healthy individuals (Floris et al., 2021). Floris et al. (2021) reported 5% of regions where infra- or supra-normals of gray matter *volume* asymmetry were more prevalent in individuals with ASD compared to healthy individuals, all case-control comparisons having small effect sizes. In a replicability analysis in an independent dataset (ABIDE) they did not report any regional differences in gray matter volume asymmetry between the ASD and healthy groups. Prior studies also found no association between handedness, IQ and regional structural brain asymmetry in ASD, SZ and healthy individuals (Kong et al., 2022). Our univariate results are in line with these findings. The multivariate results showed no relationship between the deviance of inter-regional pattern of cortical thickness asymmetry and diagnosis, indicating that normal variation in the inter-regional pattern completely encapsulates the pattern variation in ASD and SZ. It is important to note that the clustering and embedding methods used here represent just one approach to analyzing case-control variation in cortical thickness asymmetry. Other multivariate approaches, which consider a wider range of variables not focusing on brain asymmetry, have shown that multivariate clustering can successfully parse variation and identify distinct subgroups in ASD and SZ (Hong et al., 2018). The current study extends previous studies by showing, for the first time, that variation in regional asymmetry of cortical thickness in individuals with ASD and SZ is fully encapsulated by normal variation.

This study has limitations. While the present multi-center study benefited from a substantial training set, which included an Asian dataset, larger and more heterogeneous datasets, incorporating data from all continents may be important for heightened performance and validity. While multi-center studies are necessary to increase sample size it also results in including datasets lacking deep phenotyping. Hence, information about handedness, IQ and symptomatology was incomplete. However, our analysis of the relationship of handedness and IQ with asymmetry replicated previous reports, which provided additional validity for our findings. While our sample size of individuals with SZ and ASD was comparable to previous normative modeling studies, larger (multicenter) clinical samples may enable the identification of distinct clusters of patients exhibiting significant deviance (Segal et al., 2023). Finally, we focused on cortical thickness which is a property of cortical size. Other size properties such as cortical volume or novel approaches focusing on cortical shape descriptors that are independent of size, may lead to improved explanation of human variance in cortical asymmetry and may lead to improved clustering when using multivariate approaches (Chen et al., 2022).

In conclusion, variation of whole brain, regional and inter-regional asymmetry of cortical thickness in individuals with ASD was fully nested in the normative variation. For individuals with SZ, extreme deviance from the norm appeared nearly equal to that in healthy controls. Taken together, our results demonstrate that individual abnormal asymmetry of cortical thickness is generally equally present in ASD, SZ and healthy controls. Therefore our results cast doubts on asymmetry of cortical thickness as a potential biomarker for ASD and SZ.

## Data Availability

All data produced in the present work are contained in the manuscript.

