## Supplement for "Individualized cortical thickness asymmetry in Autism Spectrum Disorders and Schizophrenia"

**Content**

Supplemental Table 2	2

Supplemental Figure 1: Age distributions per dataset	3

Supplemental Figure 2: Flow diagram for inclusion/exclusion	4

Supplemental Text and Figure 3: RMSE values per region	5

Supplemental Figure 4: Distributions of normative regional z-scores for cortical thickness asymmetry	7

**Replication analyses using a higher resolution parcellation (approach A1)**

Supplemental Table 3	8

Supplemental Figure 5: RMSE values per region	9

Supplemental Text: Normative modeling-based deviance per region	10

Supplemental Figures 6 and 7: Distributions and group comparisons	12

Supplemental Text and Supplemental Figure 8: Normative modeling-based deviance per individual	17

Supplemental Figure 9: Multivariate analysis of normative modeling-based deviance	19

**Replication analyses using an alternative processing method for calculating the AI (approach A2)**

Supplemental Table 4	20

Supplemental Figure 10: RMSE values per region	21

Supplemental Text: Normative modeling-based deviance per region	22

Supplemental Figures 11 and 12: Distributions and group comparisons	24

Supplemental Text and Supplemental Figure 13 and 14: Normative modeling-based deviance per individual	26

Supplemental Figure 15: Multivariate analysis of normative modeling-based deviance	28

**Replication analyses using males only (approach A3)**

Supplemental Text: Normative modeling-based deviance per region	22

Supplemental Figures 16 and 17: Distributions and group comparisons	24

Supplemental Text and Supplemental Figure 18 and 19: Normative modeling-based deviance per individual	26

Supplemental Figure 20: Multivariate analysis of normative modeling-based deviance	28

**References**	25

Supplemental Table 2. Balanced classification accuracy scores (%) from the linear support vector machine predicting sample within healthy controls from the CV and held-out cohorts. CV, cross-validation.

| Sample | Train (CV) | Test |
| --- | --- | --- |
| ABIDE-I NYU | 0.49 | 0.50 |
| ABIDE-I USM | 0.54 | 0.66 |
| ABIDE-II GU | 0.50 | 0.53 |
| ABIDE-II OHSU | 0.49 | 0.56 |
| MITASD | 0.50 | 0.49 |
| WASHASD | 0.54 | 0.58 |
| MADRID-ASD | 0.49 | 0.52 |
| BGS | 0.55 | 0.64 |
| COBRE | 0.46 | 0.53 |
| UTRECHT | 0.58 | 0.55 |
| AOMIC-ID1000 | 0.45 | 0.51 |
| AOMIC-PIOP1 | 0.48 | 0.52 |
| AOMIC-PIOP2 | 0.49 | 0.49 |
| CAMCAN | 0.45 | 0.51 |
| DLBS | 0.45 | 0.53 |
| IXI | 0.49 | 0.50 |
| NARRATIVES | 0.48 | 0.53 |
| SALD | 0.45 | 0.50 |
| OASIS | 0.45 | 0.48 |
| ROCKLAND | 0.43 | 0.49 |

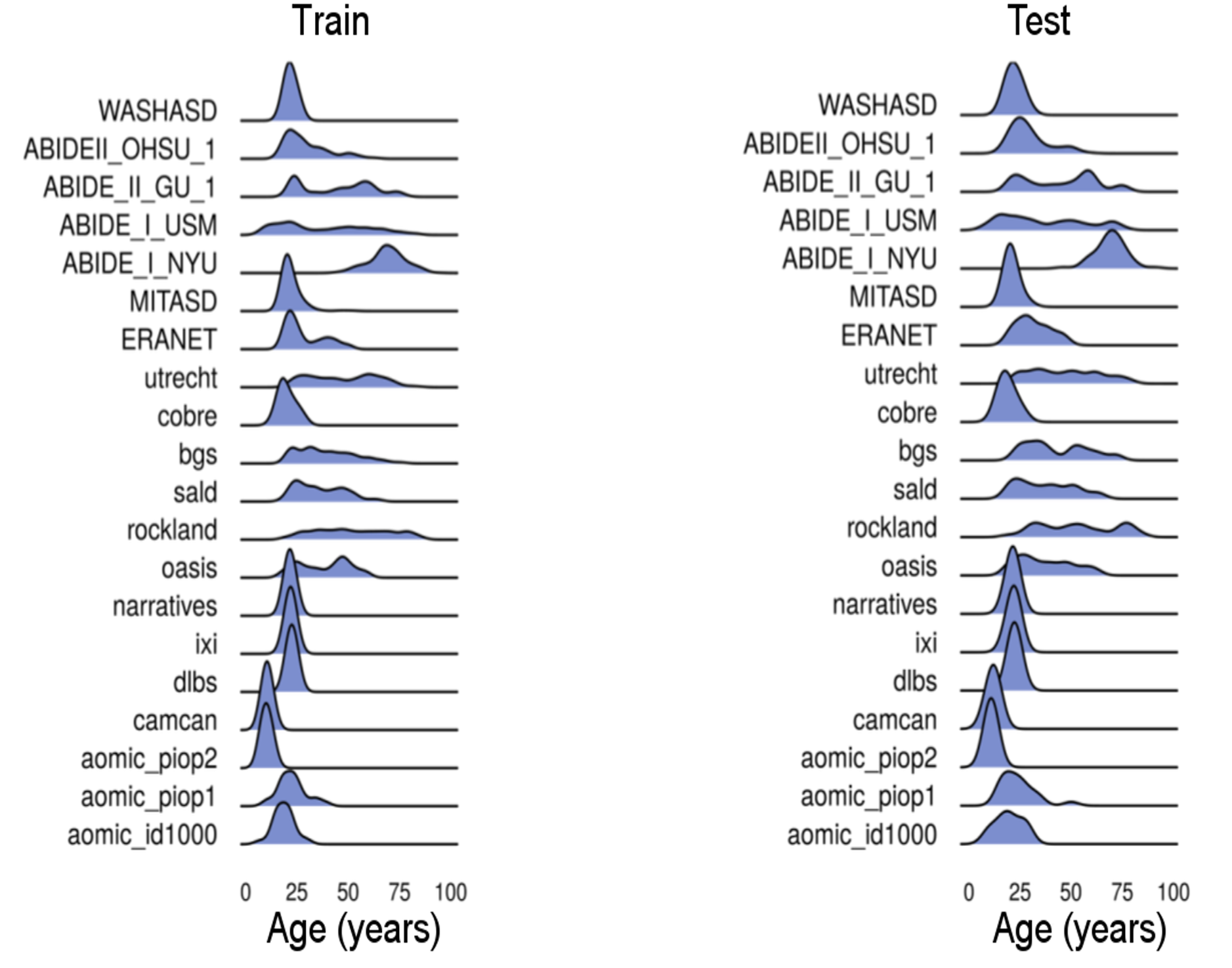

Supplemental Figure 1. The age distribution for each dataset within the training/test. For details on each dataset see Supplemental Table 1.

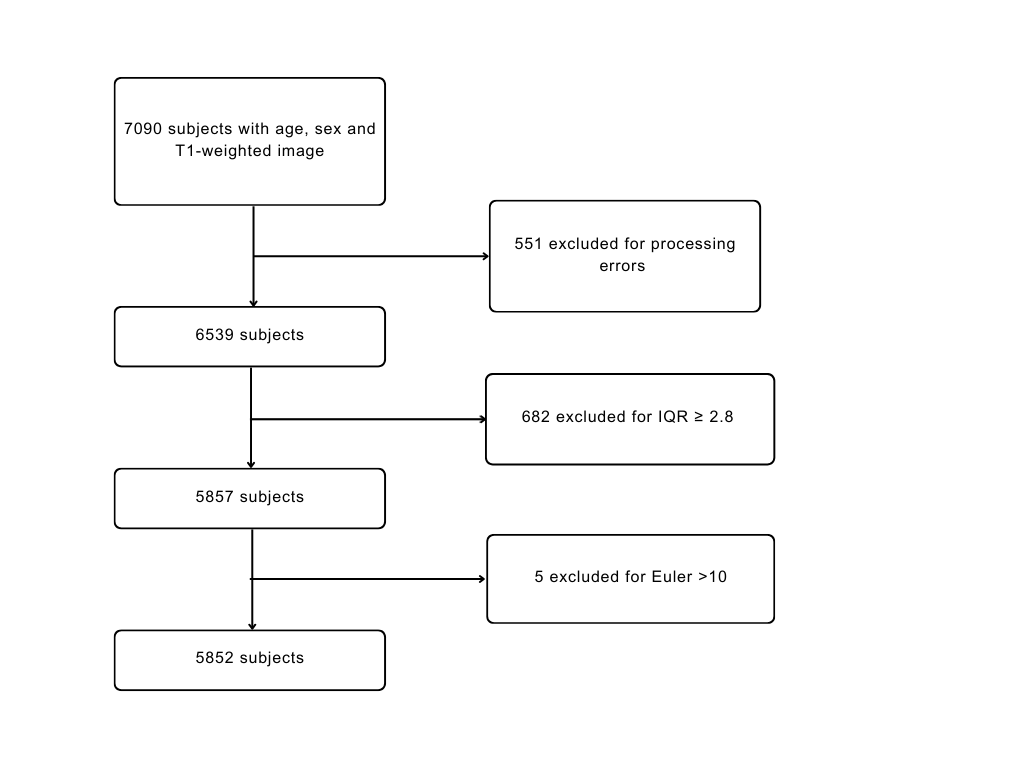

Supplemental Figure 2. Flow diagram showing the inclusion/exclusion process.

Floris et al. (2021) studied the AI of voxel-based gray matter volume and is the only study, to the best of our knowledge, to study the AI using normative modeling. The asymmetry index for each voxel was calculated using the same formula as in the current study. In the study by Floris et al. (2021) mapping was done to the right hemisphere and not to the left hemisphere. Regional RMSE values from the current study appear in general considerably lower compared to the voxel-wise RMSE values from Floris et al. (2021), see Supplemental Figure 3. The publication by Floris et al. (2021) is available at https://doi.org/10.1016/j.bpsc.2020.08.008 under a Creative Commons license available at <https://creativecommons.org/licenses/by/4.0/>.

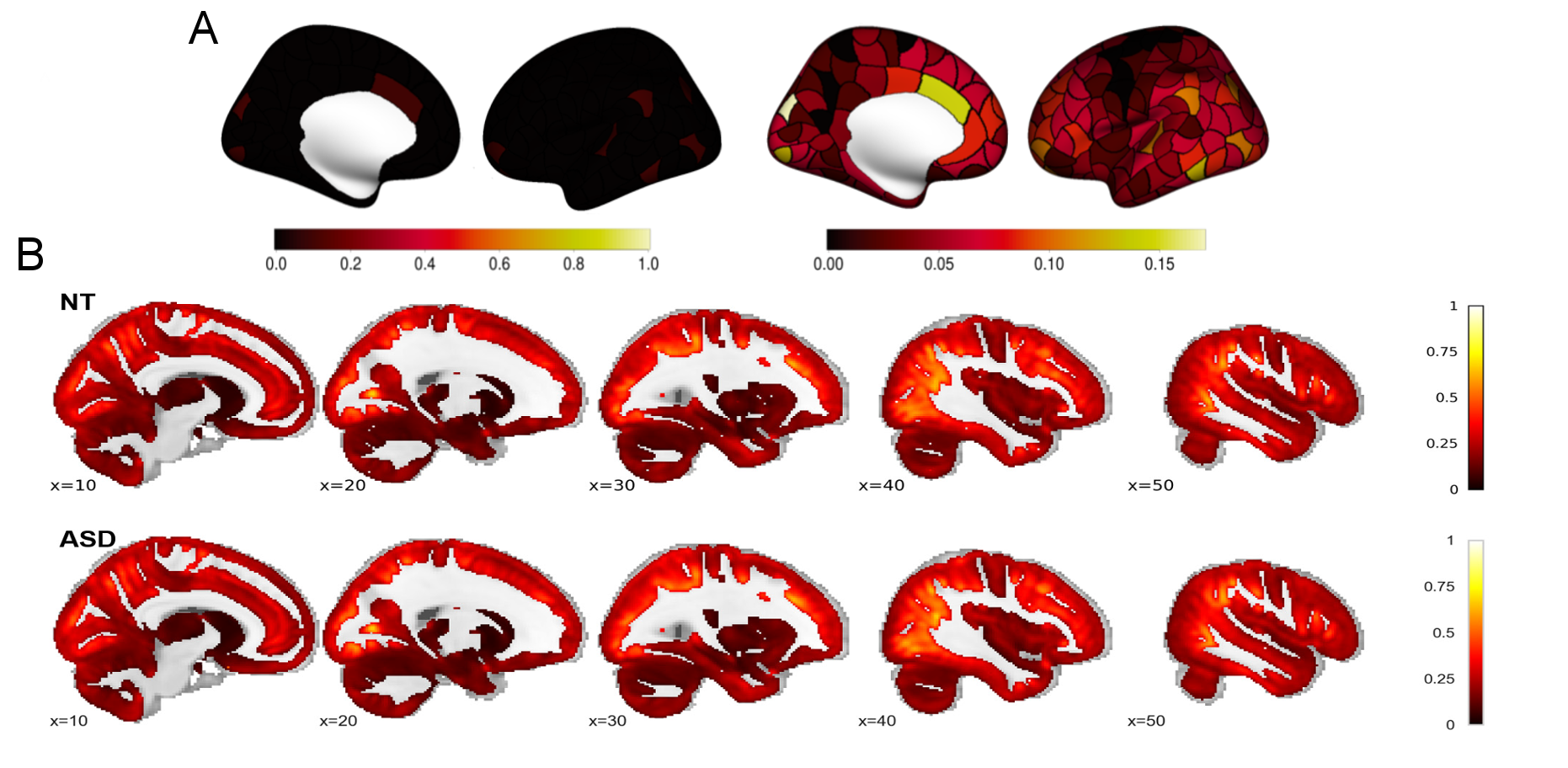

Supplemental Figure 3. **A**. Root Mean Square Error (RMSE) values of the true and predicted mean AI values are mapped to each region of the left hemisphere with the range set to (0,1) (for comparison with Floris et al., 20221) and the actual range (0,0.17). **B**. RMSE values mapped to each voxel of the right hemisphere for healthy controls (first row) and individuals with ASD (second row) in the study as depicted in Figure S4 of Floris et al. (2021). ASD, Autism Spectrum Disorders.

Supplemental Figure 4. Distribution of normative regional z-scores for cortical thickness asymmetry.

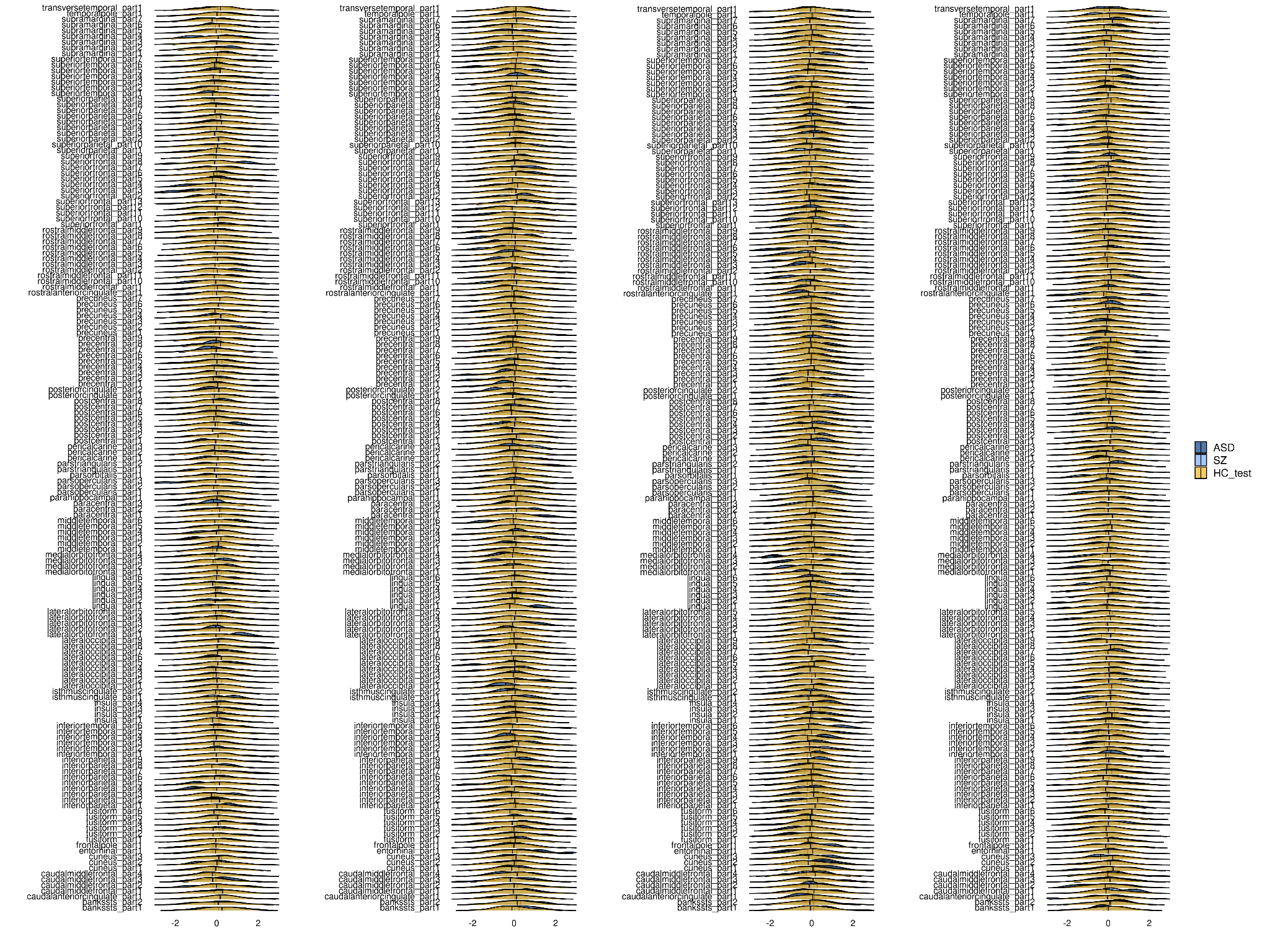

**Replication analyses using a higher resolution parcellation (A1)**

Supplemental Table 3. Balanced classification accuracy scores (%) from the linear support vector machine within healthy controls from the CV and held-out cohorts when using a higher resolution parcellation template (500 regions, Schaefer et al. 2018). CV, cross-validation.

| Sample | Train (CV) | Test |
| --- | --- | --- |
| ABIDE-I NYU | 0.50 | 0.50 |
| ABIDE-I USM | 0.50 | 0.50 |
| ABIDE-II GU | 0.50 | 0.50 |
| ABIDE-II OHSU | 0.50 | 0.54 |
| MITASD | 0.50 | 0.50 |
| WASHASD | 0.50 | 0.54 |
| MADRID-ASD | 0.50 | 0.50 |
| BGS | 0.54 | 0.59 |
| COBRE | 0.49 | 0.55 |
| UTRECHT | 0.55 | 0.62 |
| AOMIC-ID1000 | 0.49 | 0.51 |
| AOMIC-PIOP1 | 0.48 | 0.50 |
| AOMIC-PIOP2 | 0.51 | 0.50 |
| CAMCAN | 0.46 | 0.51 |
| DLBS | 0.49 | 0.50 |
| IXI | 0.50 | 0.51 |
| NARRATIVES | 0.50 | 0.51 |
| SALD | 0.52 | 0.51 |
| OASIS | 0.48 | 0.51 |
| ROCKLAND | 0.48 | 0.49 |

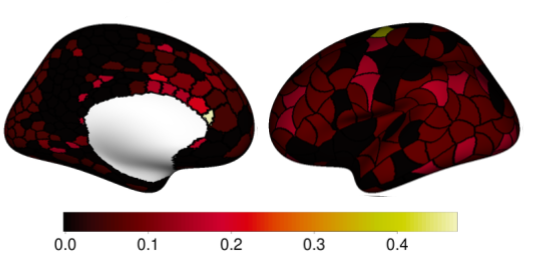

Supplemental Figure 5. Root Mean Square Error (RMSE) values of the true and predicted mean AI values when using a higher resolution parcellation for calculating the AI are mapped to each region of the left hemisphere. AI, Asymmetry Index.

#### *Deviance per region*

The percentages of each group for each region and whether the group difference was significant are included in Supplementary Data. The distributions of the z-scores of the ASD and SZ groups overlapped with HC_test_, see Supplemental Figure 10. The infra-normal regional AI z-scores were noted in 0% to 8.15% of individuals with ASD, 0% to 10.8% of individuals with SZ and 0.57% to 5.89% of HC_test_; the corresponding ranges for supra-normal z-scores were 0% to 8.15% in individuals with ASD, 0.35% to 8.36% in SZ, and 0.76% to 5.7% in HC_test_. There were no significant group differences in the regional proportion of infra-normals or supra-normals between HC_test_ and ASD. Out of 500 regions there were twelve regions where the proportion of infra-normals differed significantly between HC_test_ and SZ, see Supplemental Figure 11. In all of these regions the SZ group had a higher percentage of infra-normals compared to the HC_test_ group, the Cohen’s ω ranged from 0.11 to 0.18. Regions were not clustered but located mainly across the inferior, medial and superior frontal cortex. In all twelve regions, the groups of infra-normals of HC_test_ and SZ had rightward asymmetry and for all regions there was no significant difference in the mean AI between the groups of infra-normals of HC_test_ and SZ, the Cohen’s d values ranged from -0.42 to 0.3. There was one region in the inferior frontal cortex where the proportion of supra-normals differed significantly between HC_test_ and SZ (HC_test_ = 2.09%, SZ = 8.36%, 𝞆^2^ = 17.7, P_FDR_< 0.05, ω = 0.15). The groups of supra-normals of HC_test_ and SZ both had leftward asymmetry in that region and they did not differ from each other (mean AI supra-normals HC_test_ = 0.30, mean AI supra-normals SZ = 0.28, t= -0.81, P > 0.05, Cohen’s d: -0.29).

Infra-normal z-scores in any regional AI were observed in 100% for all the groups; the corresponding supra-normal z-scores were 100% for ASD and SZ, and 99.8% for HC_test_.

Supplemental Figure 6. Distribution of normative regional z-scores for cortical thickness asymmetry (4 pages).

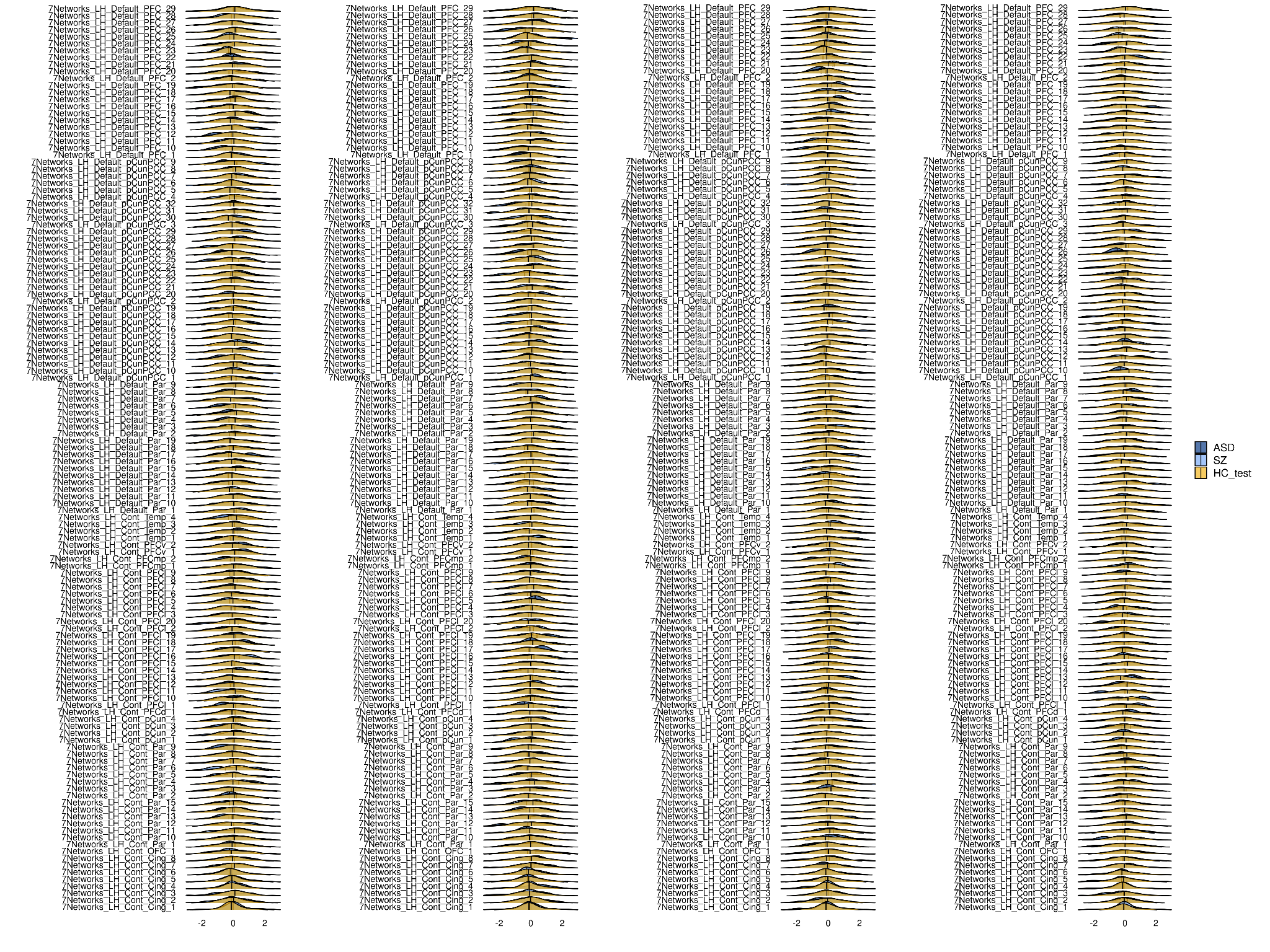

Supplemental figure 6. Continuation.

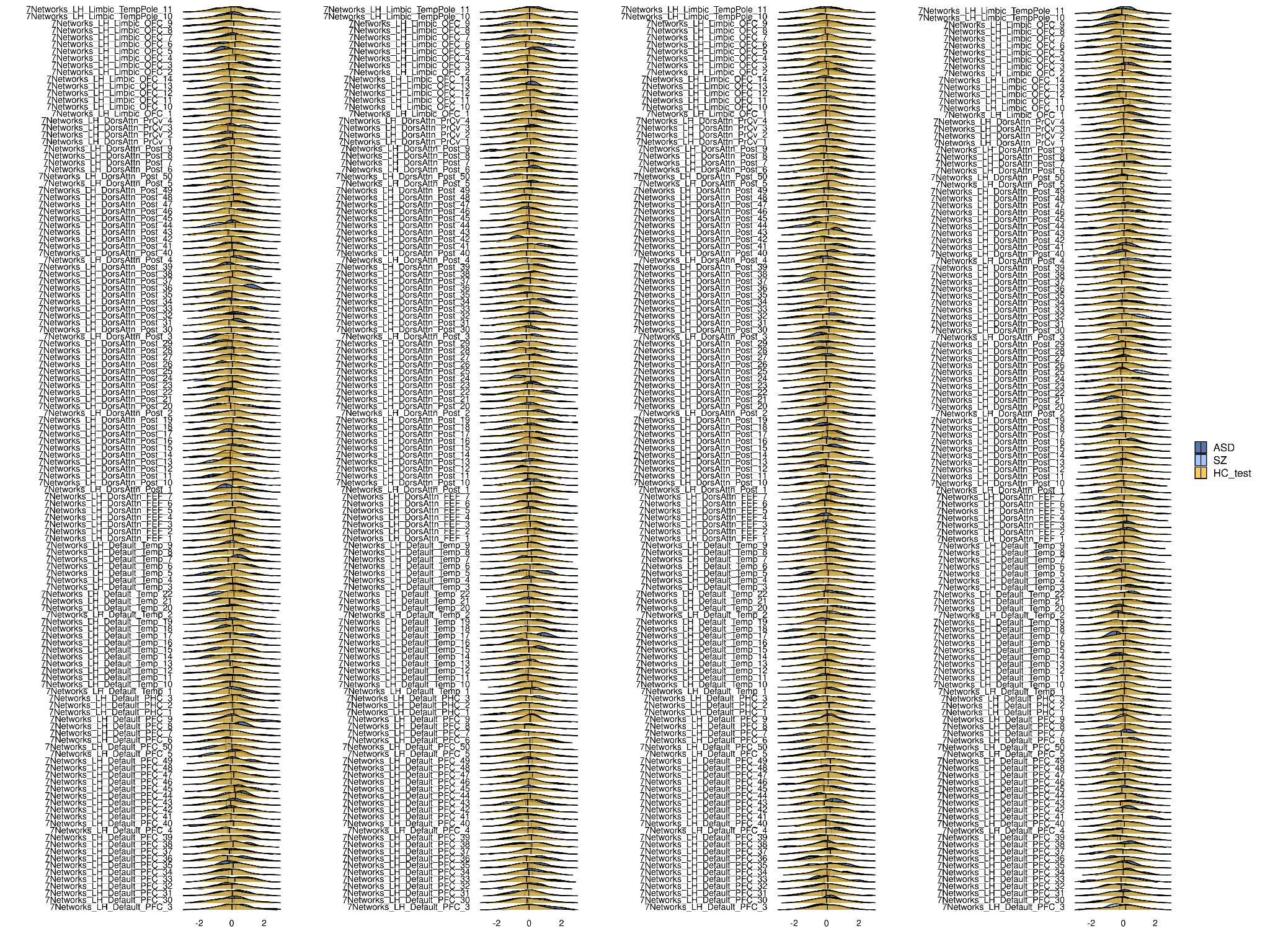

Supplemental figure 6. Continuation.

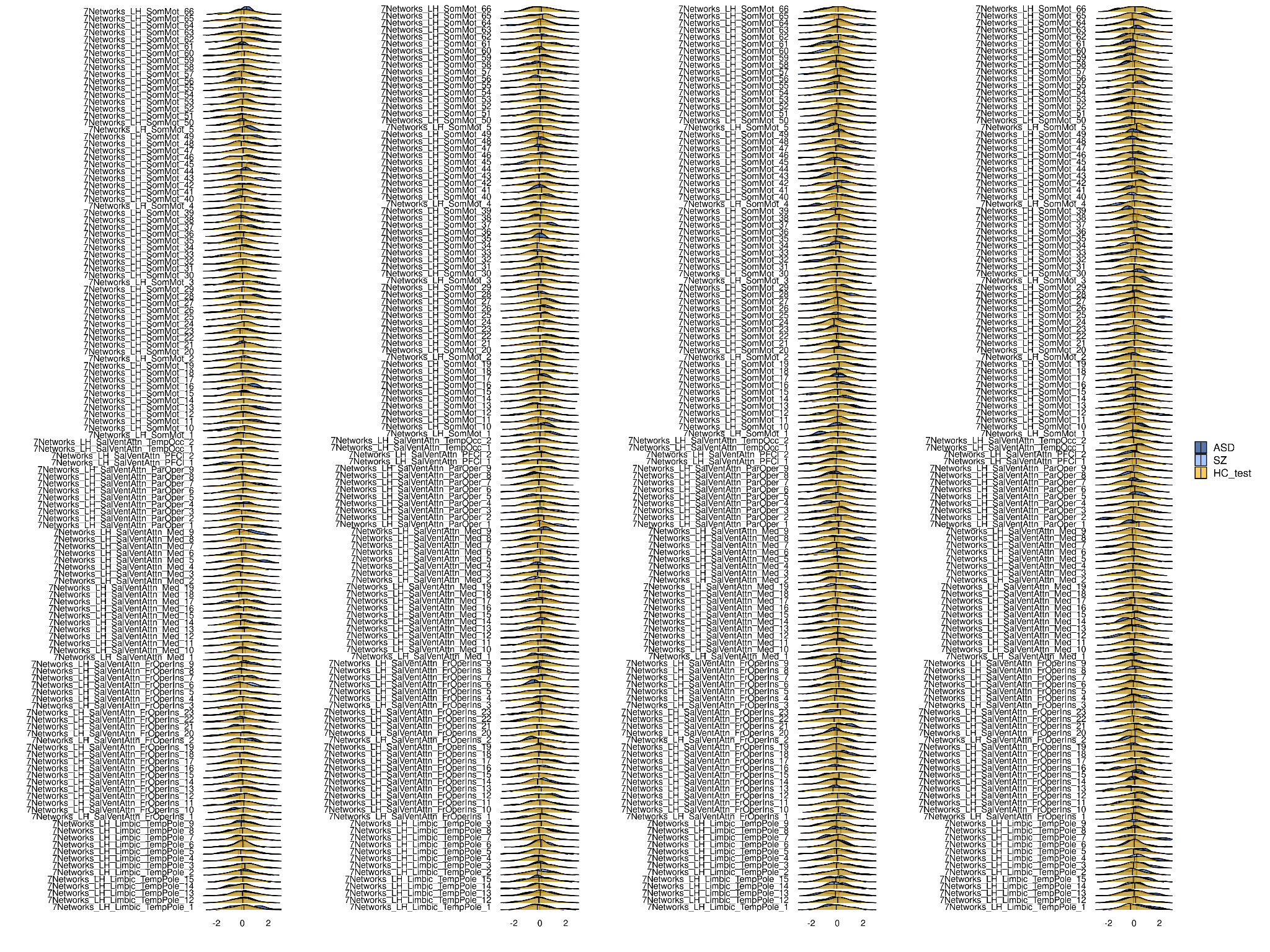

Supplemental figure 6. Continuation.

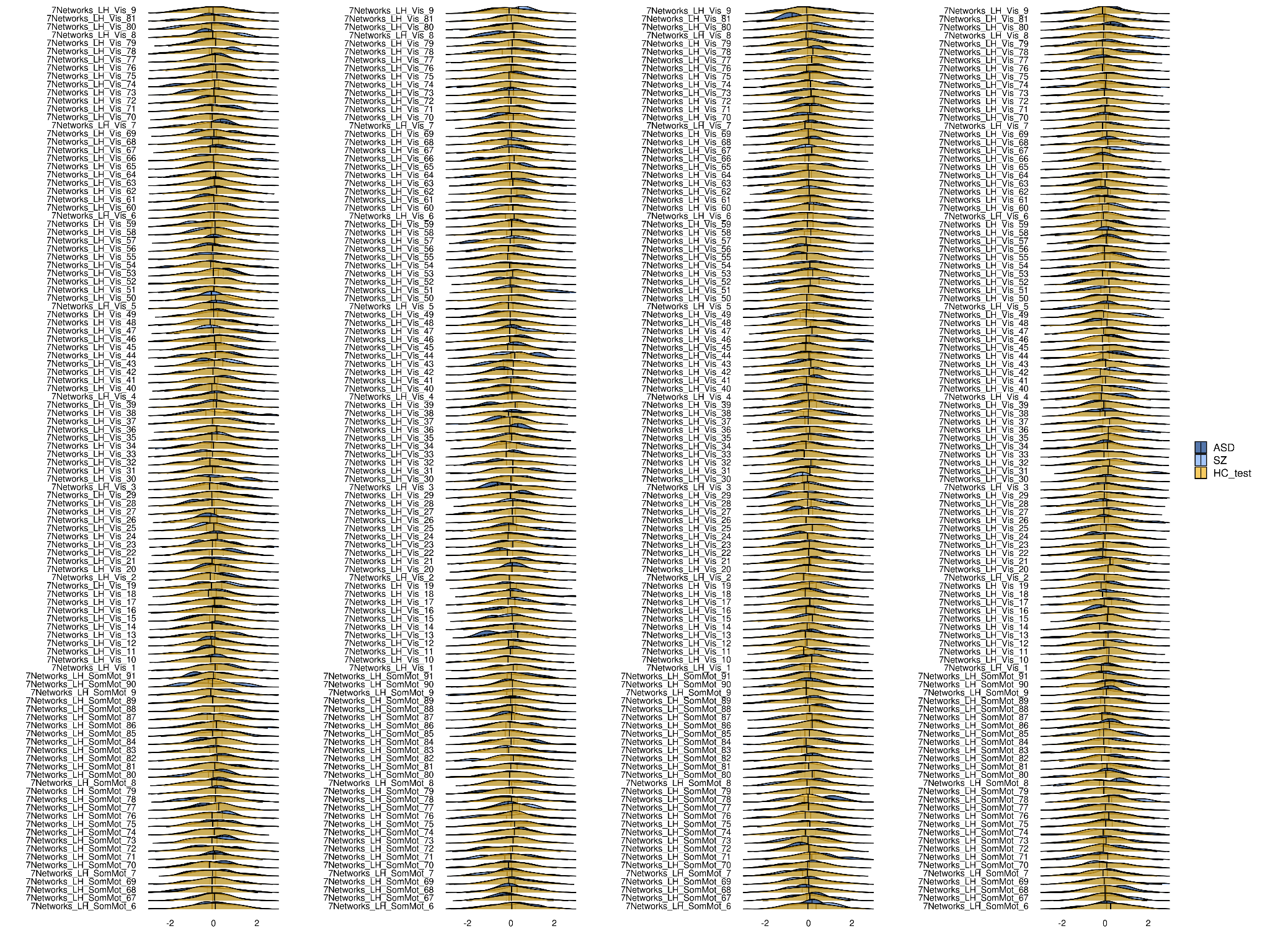

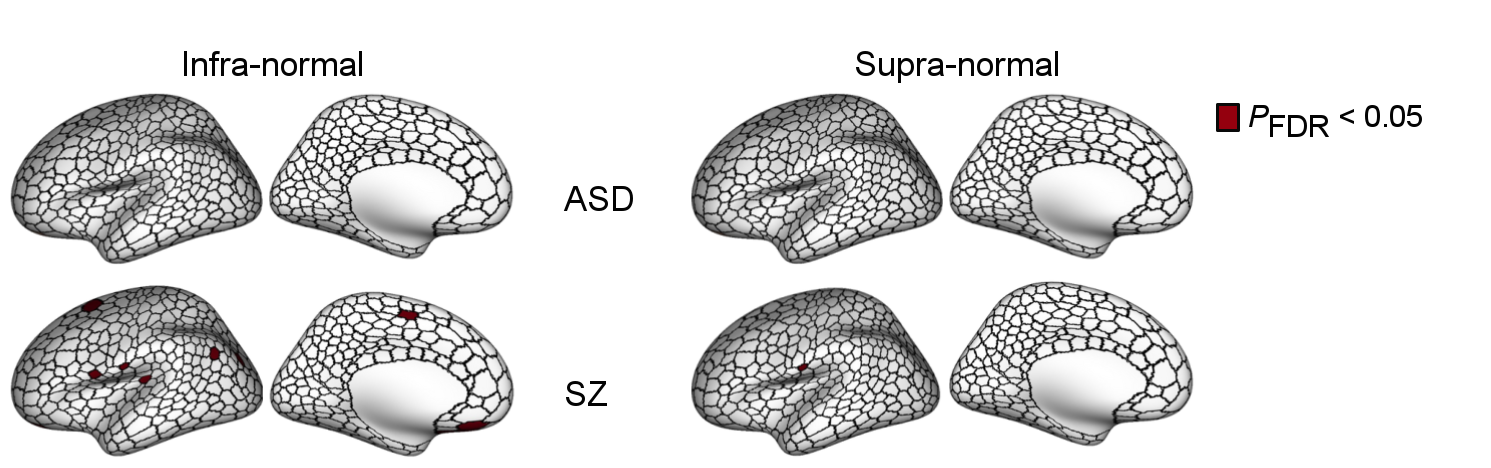

Supplemental Figure 7. **A**. Case-control comparisons using a higher resolution parcellation (500 regions, Schaefer et al. 2018). We compared each region of the percentage map from HC_test_ and each clinical group’s percentage map using permutation chi-square tests yielding a map showing regions with significantly different percentage of infra- and supra-normal AI deviations in cases compared with controls. HC_test_, healthy individuals from the test set; ASD, Autism Spectrum Disorders; SZ, schizophrenia. Data used to generate this figure can be found in Supplementary Data.

#### *Deviance of regional AI per individual*

The distributions of the proportion of individuals with regional infra- and supra-normal deviance were similar for ASD and HC_test_; there were no group differences in the average number of infra- and supra-normal regions between ASD and HC_test_, see Supplementary Figure 8. The SZ group had a higher average number of infra- and supra-normal regions compared to HC_test_ (infra-normal: z=-5.98, p<0.01 , supra-normal: z=-5.44, p<0.01), see Supplementary Figure 8.

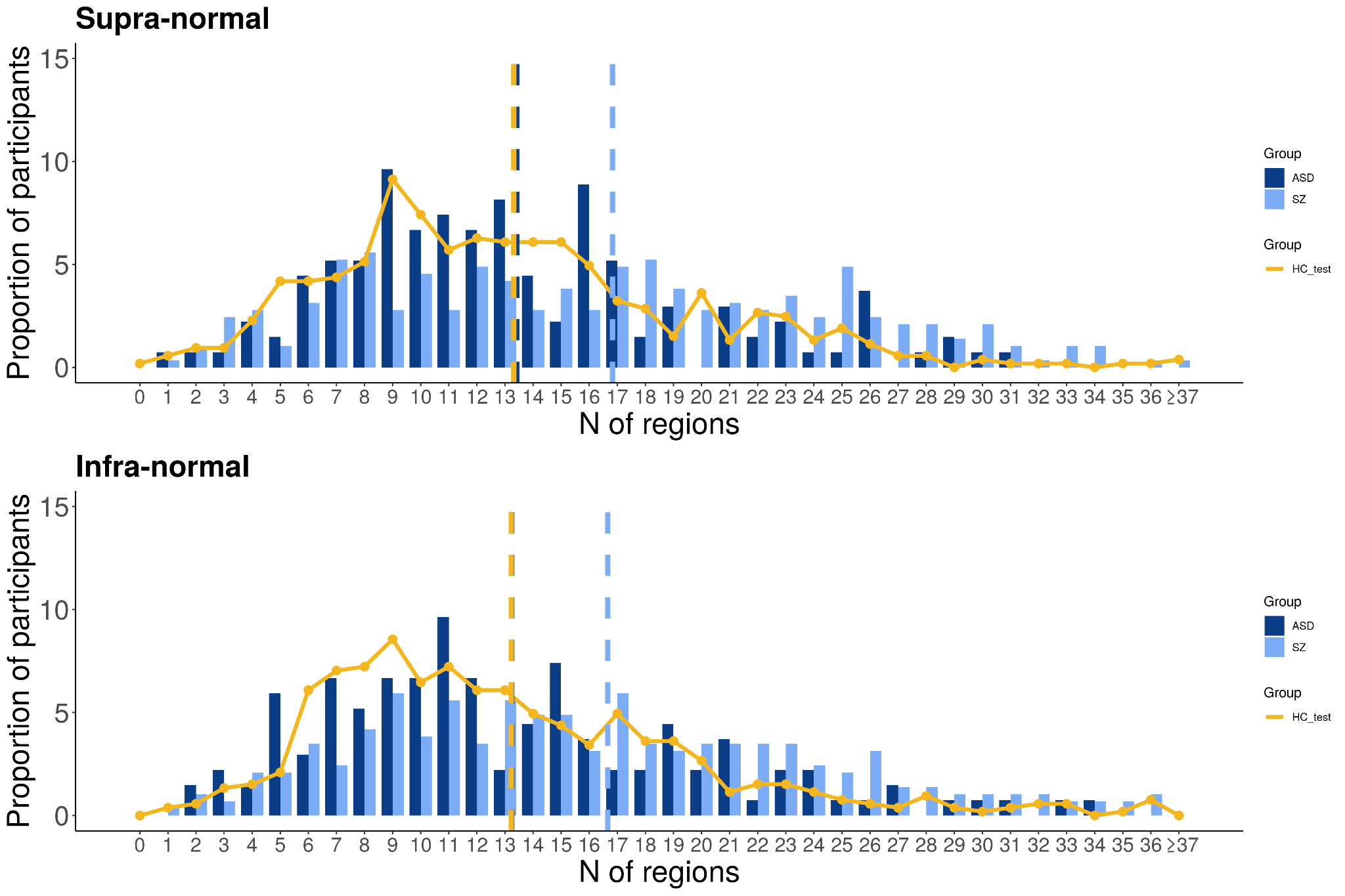

Supplemental Figure 8. Distribution of the number of regions with infra- or supra-normal deviance per individual. Bar plots and curves display the distribution of the proportion of individuals per amount of regions with supra-normal and infra-normal deviations. HC_test_, healthy individuals from the test set; ASD, Autism Spectrum Disorders; SZ, schizophrenia.

*Multivariate analysis*

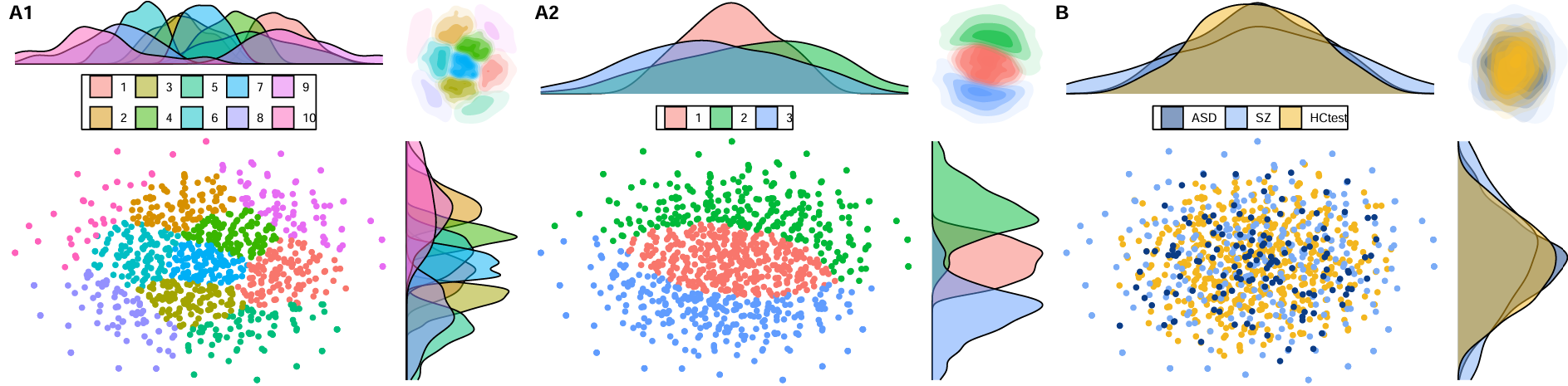

Supplemental Figure 9. Panels A1 and A2 present the optimum (A1) and three cluster results (A2) of k-medoid clustering applied to the 2D embedding of z-scores for regional cortical thickness AI, generated using tSNE. Panel B demonstrates that these clusters did not yield any meaningful differentiation based on diagnosis. AI, asymmetry index; HC_test_, healthy individuals from the test set; ASD, Autism Spectrum Disorders; SZ, schizophrenia.

**Replication analyses using an alternative processing method for calculating the AI (A2)**

Supplemental Table 4. Balanced classification accuracy scores (%) from the linear support vector machine predicting sample within healthy controls from the CV and held-out cohorts when using an alternative image processing pipeline for calculating the AI. CV, cross-validation.

| Sample | Train (CV) | Test |
| --- | --- | --- |
| ABIDE-I NYU | 0.49 | 0.50 |
| ABIDE-I USM | 0.50 | 0.59 |
| ABIDE-II GU | 0.50 | 0.54 |
| ABIDE-II OHSU | 0.48 | 0.52 |
| MITASD | 0.50 | 0.54 |
| WASHASD | 0.50 | 0.61 |
| MADRID-ASD | 0.49 | 0.56 |
| BGS | 0.52 | 0.65 |
| COBRE | 0.46 | 0.56 |
| UTRECHT | 0.58 | 0.56 |
| AOMIC-ID1000 | 0.45 | 0.51 |
| AOMIC-PIOP1 | 0.45 | 0.50 |
| AOMIC-PIOP2 | 0.51 | 0.50 |
| CAMCAN | 0.44 | 0.51 |
| DLBS | 0.49 | 0.50 |
| IXI | 0.50 | 0.54 |
| NARRATIVES | 0.49 | 0.48 |
| SALD | 0.48 | 0.56 |
| OASIS | 0.45 | 0.49 |
| ROCKLAND | 0.44 | 0.50 |

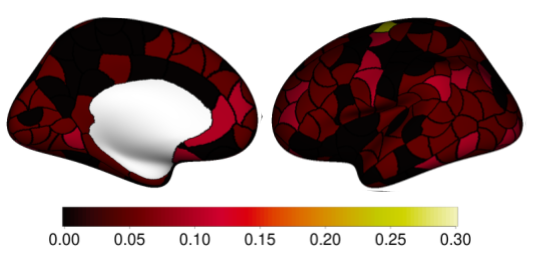

Supplemental Figure 10. Root Mean Square Error (RMSE) values of the true and predicted mean AI values when using an alternative image processing pipeline for calculating the AI are mapped to each region of the left hemisphere. AI, Asymmetry Index.

#### *Deviance per region*

The percentages of each group for each region and whether the group difference was significant are included in the Supplementary Data. The distributions of the z-scores of the ASD and SZ groups overlapped with HC_test_, see Supplemental Figure 6. The infra-normal regional AI z-scores were noted in 0% to 6.67% of individuals with ASD, 0% to 8.71% of individuals with SZ and 1.3% to 5.5% of HC_test_; the corresponding ranges for supra-normal z-scores were 0% to 8.15% in individuals with ASD, 0.67% to 7.67% in SZ, and 1.14% to 4.75% in HC_test_. There were no significant group differences in the regional proportion of infra-normals or supra-normals between HC_test_ and ASD. Out of 160 regions, there was one region in the medial orbitofrontal cortex where the proportion of infra-normals differed significantly between HC_test_ and SZ (HC_test_ = 1.52%, SZ = 6.62%, 𝞆^2^ = 15.04, P_FDR_< 0.05, Cohen’s ω (ω) = 0.14), see Supplemental Figure 7. The infra-normals groups of HC_test_ and SZ both had rightward asymmetry in that region in the medial orbitofrontal cortex and they did not differ from each other (mean AI infra-normals HC_test_ = -0.17, mean AI infra-normals SZ = -0.21, t= -1.60, P > 0.05, d = -0.48 ). There was one region in the fusiform gyrus where the proportion of supra-normals differed significantly between HC_test_ and SZ (HC_test_ = 2.28%, SZ = 7.67%, 𝞆^2^ = 13.43, P_FDR_< 0.05, ω = 0.13). The supra-normals groups of HC_test_ and SZ both had leftward asymmetry in that region in the fusiform gyrus and they did not differ from each other (mean AI supra-normals HC_test_ = 0.16, mean AI supra-normals SZ = 0.15, t= -0.16, P > 0.05, d = -0.05 ). Infra-normal z-scores in any regional AI were observed in 94.07%, 95.47% and 95.06% across ASD, SZ, and HC_test_, respectively; the corresponding supra-normal z-scores were 100% for all groups.

Supplemental Figure 11. Distribution of normative regional z-scores for cortical thickness asymmetry.

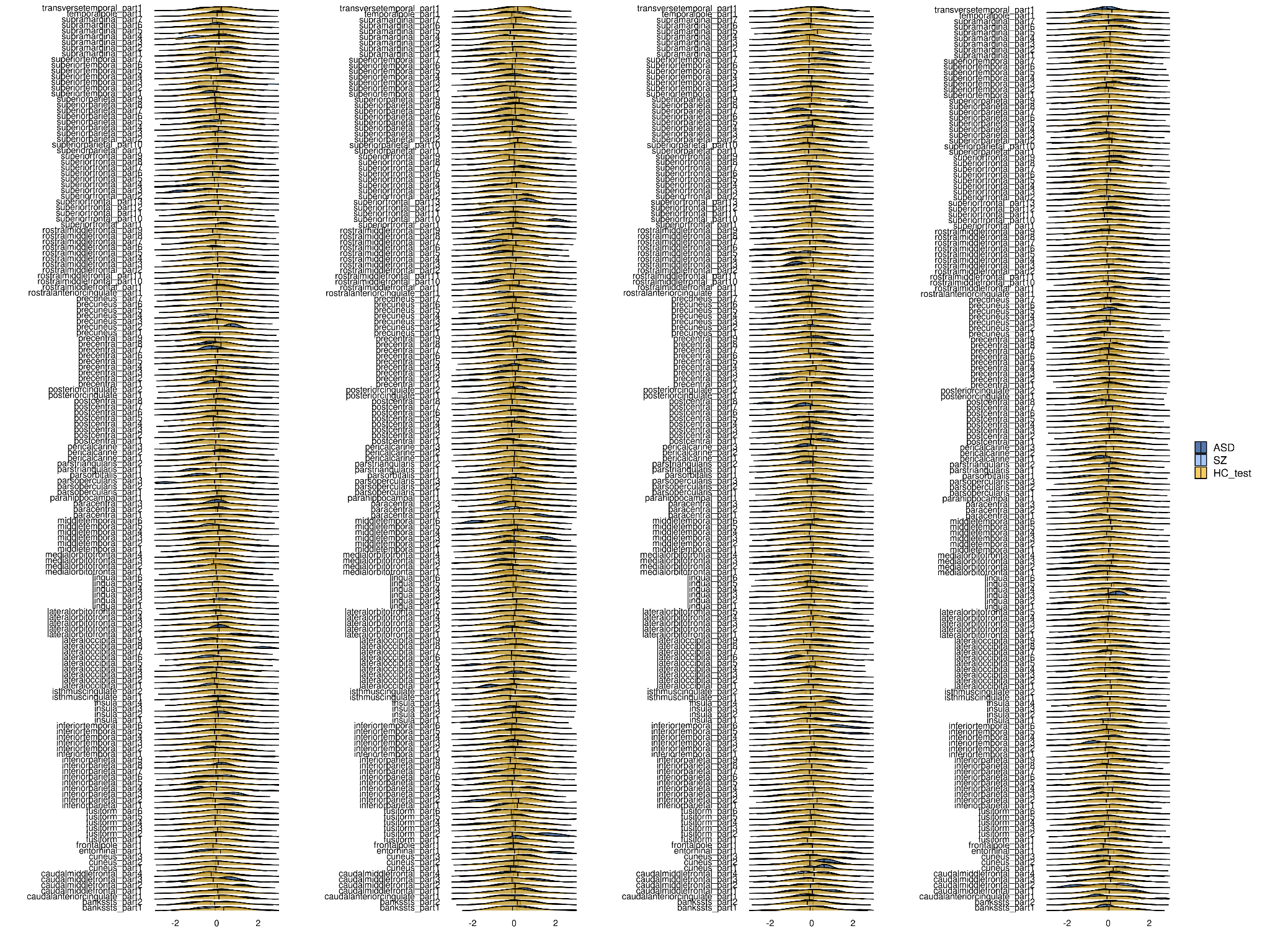

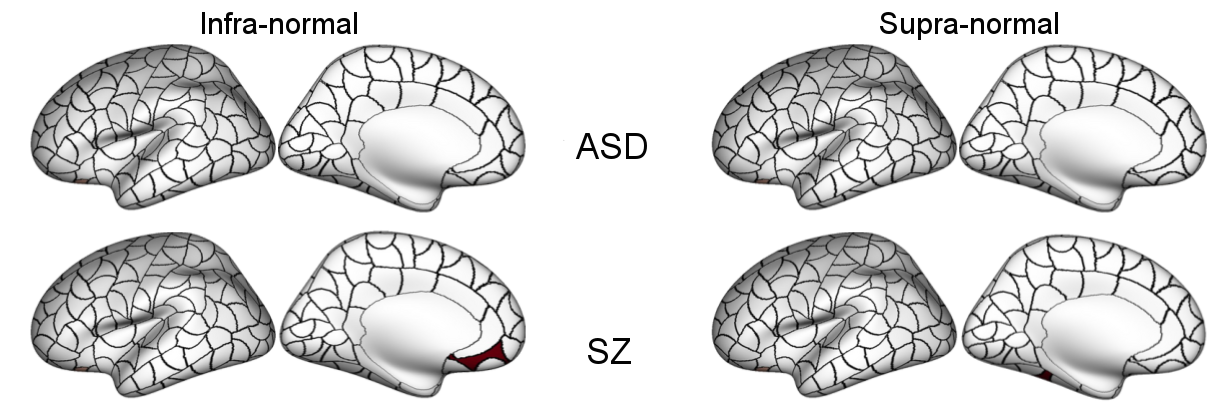

Supplemental Figure 12. **A**. Case-control comparisons using an alternative processing pipeline for calculating the AI. We compared each region of the percentage map from HC_test_ and each clinical group’s percentage map using permutation chi-square tests yielding a map showing regions with significantly different percentage of infra- and supra-normal AI deviations in cases compared with controls. HC_test_, healthy individuals; ASD, Autism Spectrum Disorders; SZ, schizophrenia. Data used to generate this figure can be found in Supplementary Data.

*Whole brain*

Here we calculated the z-scores for the whole brain AI and counted the number of individuals who had infra- or supra-normal deviance. The distributions of the z-scores of the whole brain AI overlapped for HC_test_, ASD, and SZ groups, see Supplemental Figure 13. There were no differences in z-scores for whole brain AI between HC_test_ and ASD (P > 0.05, Cohen’s d (d) = -0.1) and between HC_test_ and SZ (P > 0.05, d = -0.06). There were no differences in the proportion of individuals with infra-normal or supra-normal deviance between HC_test_ and ASD for whole brain AI (proportion of individuals with infra-normal z-scores: HC_test_ = 2.66%, ASD = 1.48%, P > 0.05, Cohen’s ω (ω) = 0.06; proportion of individuals with supra-normal z-scores: HC_test_ = 2.47%, ASD = 5.93%, P > 0.05, ω = 0.15) and between HC_test_ and SZ (proportion of individuals with infra-normal z-scores: SZ = 2.43%, P > 0.05, ω = 0.01; proportion of individuals with supra-normal z-scores: SZ = 2.09%, P > 0.05, ω = 0.02).

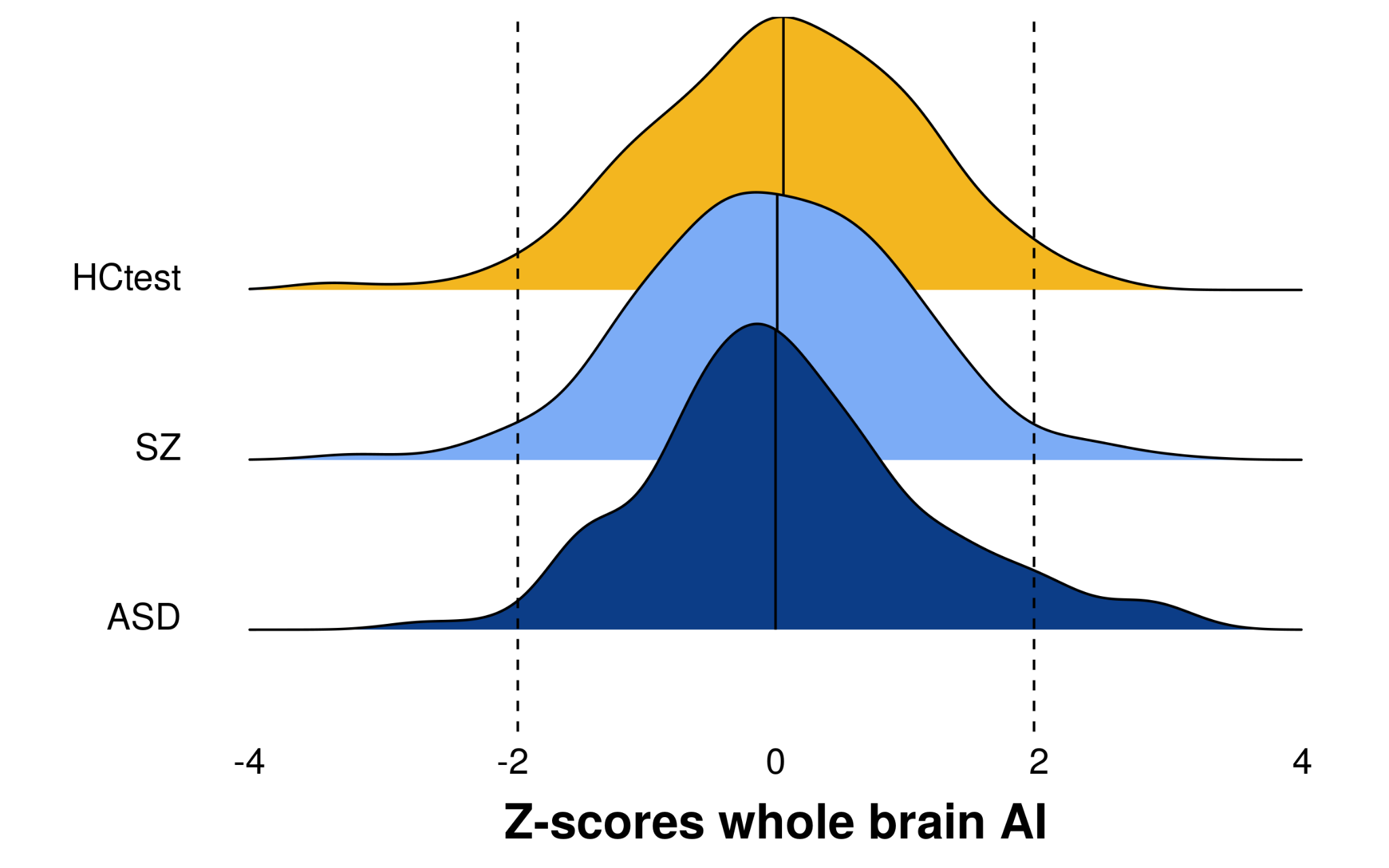

Supplemental Figure 13. The distributions of the normative modeling based z-scores for the whole brain AI in HC_test_, SZ, and ASD. The vertical line represents the median for each group. The dotted lines represent |z| = 1.96. HC_test_, healthy controls from the test set; ASD, Autism Spectrum Disorders; SZ, schizophrenia.

#### *Deviance of regional AI per individual*

The distributions of the proportion of individuals with regional infra- and supra-normal deviance were similar for ASD and HC_test_; there were no group differences in the average number of infra- and supra-normal regions between ASD and HC_test_, see Supplementary Figure 14. The SZ group had a higher average number of infra- and supra-normal regions compared to HC_test_ (infra-normal: z=-4.94, p<0.01 , supra-normal: z=-5.67, p<0.01), see Supplementary Figure 14.

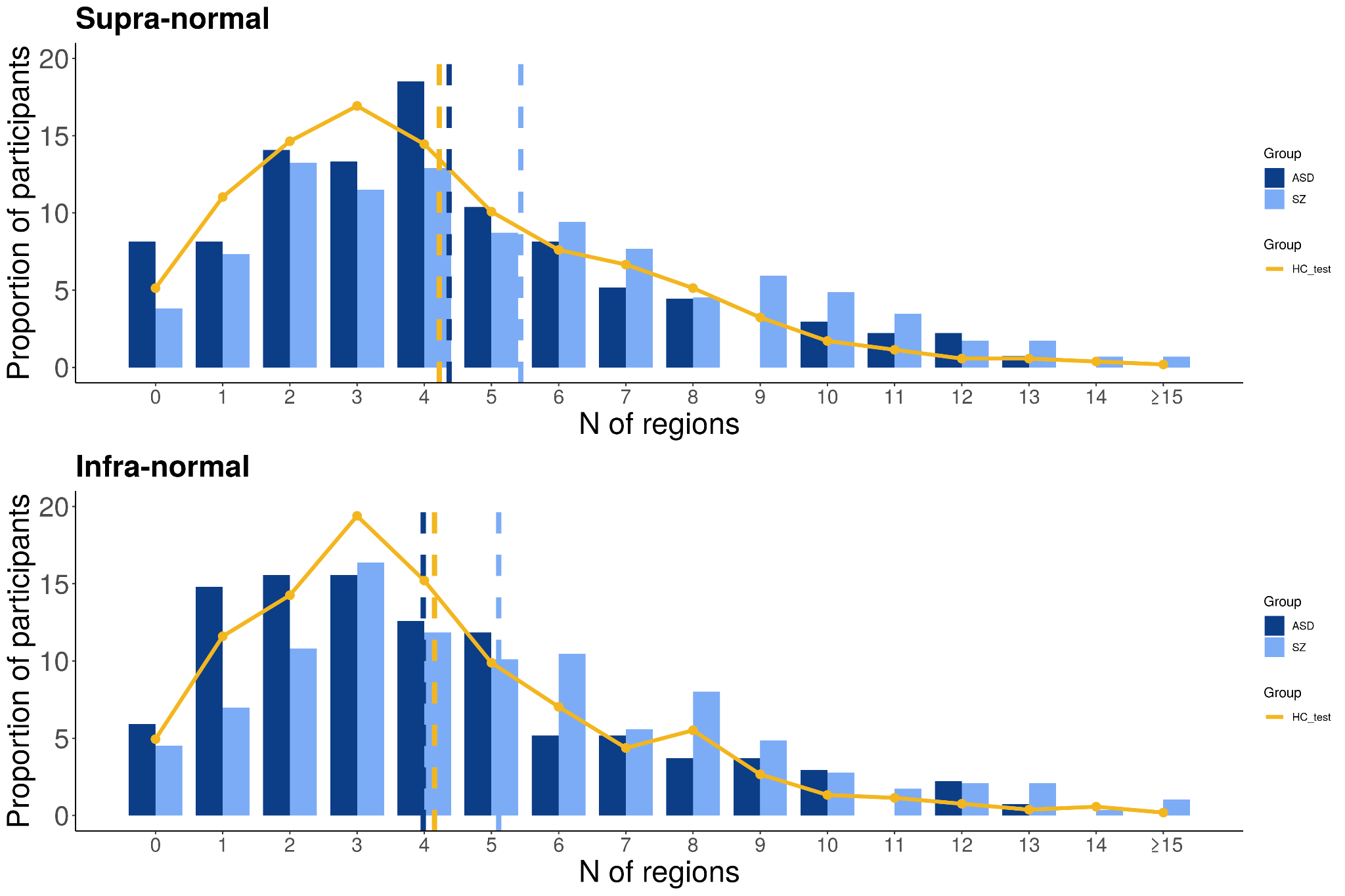

Supplemental Figure 14. Distribution of the number of regions with infra- or supra-normal deviance per individual. Bar plots and curves display the distribution of the proportion of individuals per amount of regions with supra-normal and infra-normal deviations. HC_test_, healthy individuals from the test set; ASD, Autism Spectrum Disorders; SZ, schizophrenia.

*Multivariate analysis*

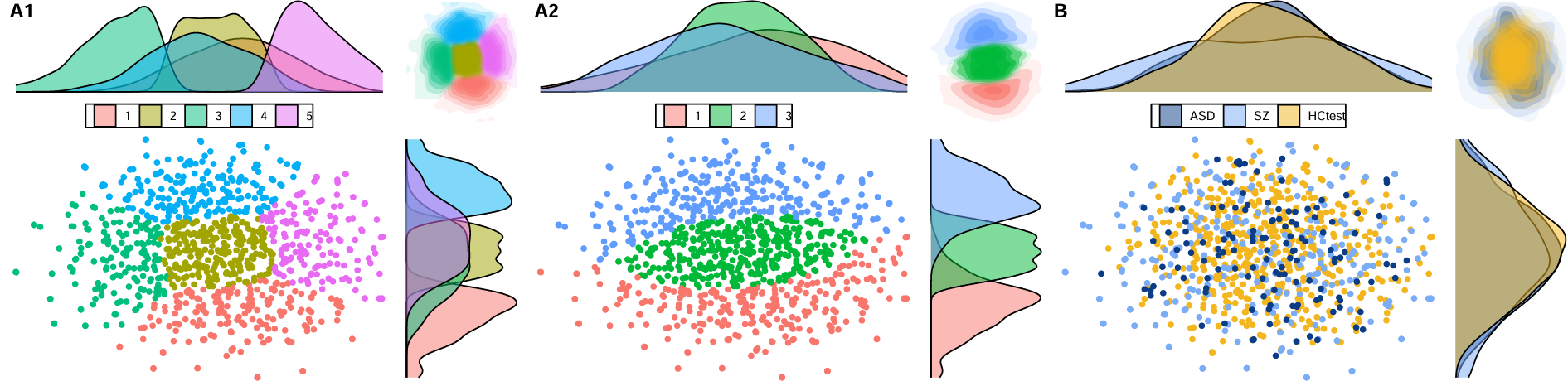

Supplemental Figure 15. Panels A1 and A2 present the optimum (A1) and three cluster results (A2) of k-medoid clustering applied to the 2D embedding of z-scores for regional cortical thickness AI, generated using tSNE. Panel B demonstrates that these clusters did not yield any meaningful differentiation based on diagnosis. AI, Asymmetry Index; HC_test_, healthy individuals from the test set; ASD, Autism Spectrum Disorders; SZ, schizophrenia.

**Confirmatory analyses using males only (A3)**

#### *Deviance per region*

The percentages of each group for each region and whether the group difference was significant are included in Supplementary Data. The distributions of the z-scores of the ASD and SZ groups overlapped with HC_test_, see Supplemental Figure 16. The infra-normal regional AI z-scores were noted in 0% to 7.14% of individuals with ASD, 0% to 7.33% of individuals with SZ and 0.47% to 6.13% of HC_test_; the corresponding ranges for supra-normal z-scores were 0% to 10.7% in individuals with ASD, 0.43% to 7.76% in SZ, and 0% to 5.19% in HC_test_. There were no significant group differences in the regional proportion of infra-normals or supra-normals between HC_test_ and ASD, nor between HC_test_ and SZ, see Supplemental Figure 17. Infra-normal z-scores in any regional AI were observed in 95% for ASD, 96% for SZ and 93% for HC_tes_; the corresponding supra-normal z-scores were 94% for ASD, 96% for SZ, and 95% for HC_test_.

Supplemental Figure 16. Distribution of normative regional z-scores for cortical thickness asymmetry.

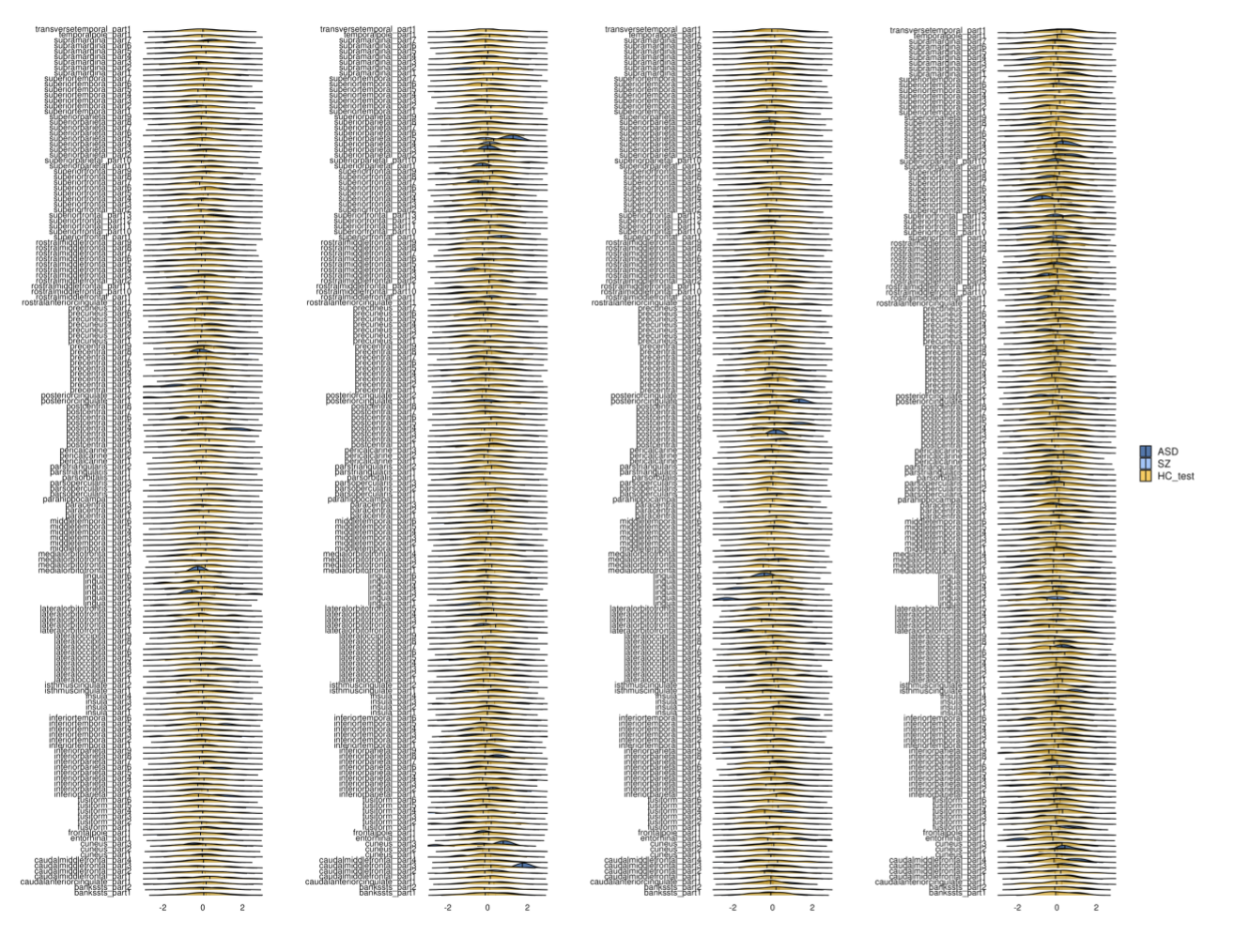

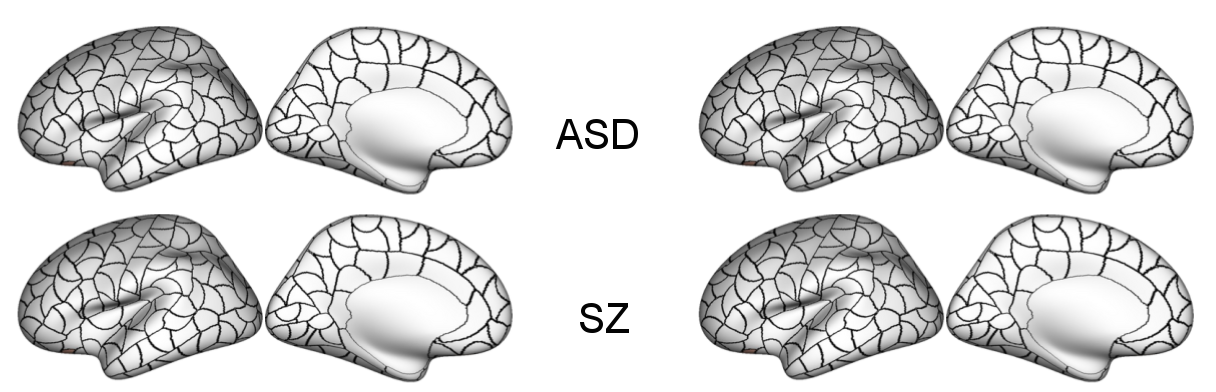
Supplemental Figure 17. **A**. Case-control comparisons using only males. We compared each region of the percentage map from HC_test_ and each clinical group’s percentage map using permutation chi-square tests yielding a map showing regions with significantly different percentage of infra- and supra-normal AI deviations in cases compared with controls. HC_test_, healthy individuals; ASD, Autism Spectrum Disorders; SZ, schizophrenia. Data used to generate this figure can be found in Supplementary Data.

*Global deviance per individual*

The distributions of the global deviance z-scores overlapped for HC_test_, ASD and SZ groups, see Supplemental Figure 18. There were no significant group differences in average global deviance z-scores between HC_test_ and ASD (P > 0.05, d = -0.01) and between HC_test_ and SZ (P > 0.05, d = -0.07). There were no significant group differences for global deviance in the proportion of infra-normals or supra-normals between HC_test_ and ASD (infra-normals: HC_test_ = 1.42%, ASD = 1.79%, P > 0.05, ω = 0.03; supra-normals: HC_test_ = 2.83%, ASD = 7.14%, P > 0.05, ω = 0.20) and between HC_test_ and SZ (infra-normals: SZ = 3.02%, P > 0.05, ω = 0.11; supra-normals: SZ = 3.88%, P > 0.05, ω = 0.06).

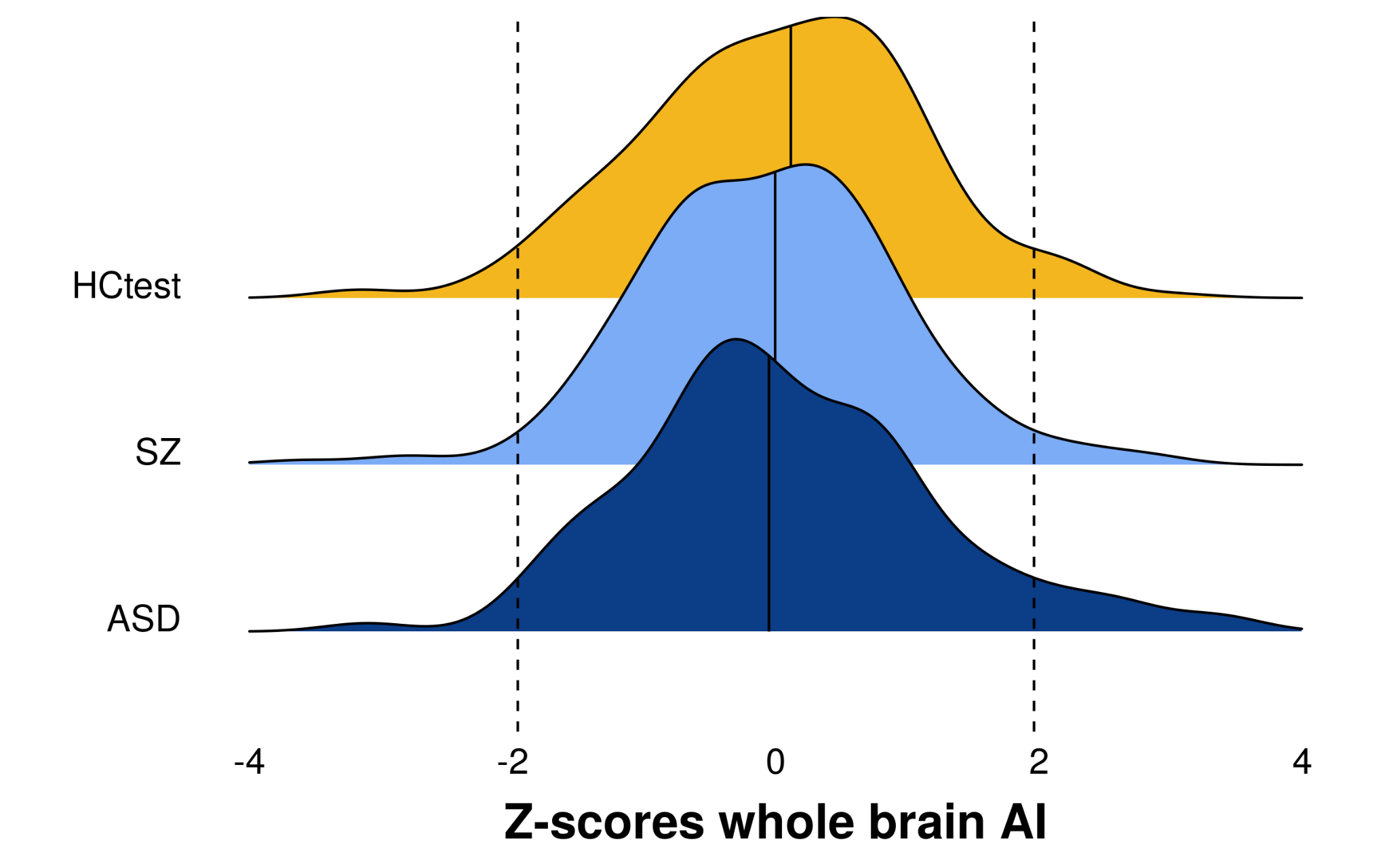

Supplemental Figure 18. The distributions of the global deviance z-scores in only males of HC_test_, SZ and ASD. The vertical line represents the median. The dotted lines represent z = |1.96|. HC_test_, healthy individuals from the test set; ASD, Autism Spectrum Disorders; SZ, schizophrenia.

#### *Deviance of regional AI per individual*

The distributions of the proportion of individuals with regional infra- and supra-normal deviance were similar for ASD and HC_test_; there were no group differences in the average number of infra- and supra-normal regions between ASD and HC_test_, see Supplemental Figure 19. The SZ group had a higher average number of infra- and supra-normal regions compared to HC_test_ (infra-normal: z=-3.32, p<0.01 , supra-normal: z=-3.54, p<0.01), see Supplemental Figure 19.

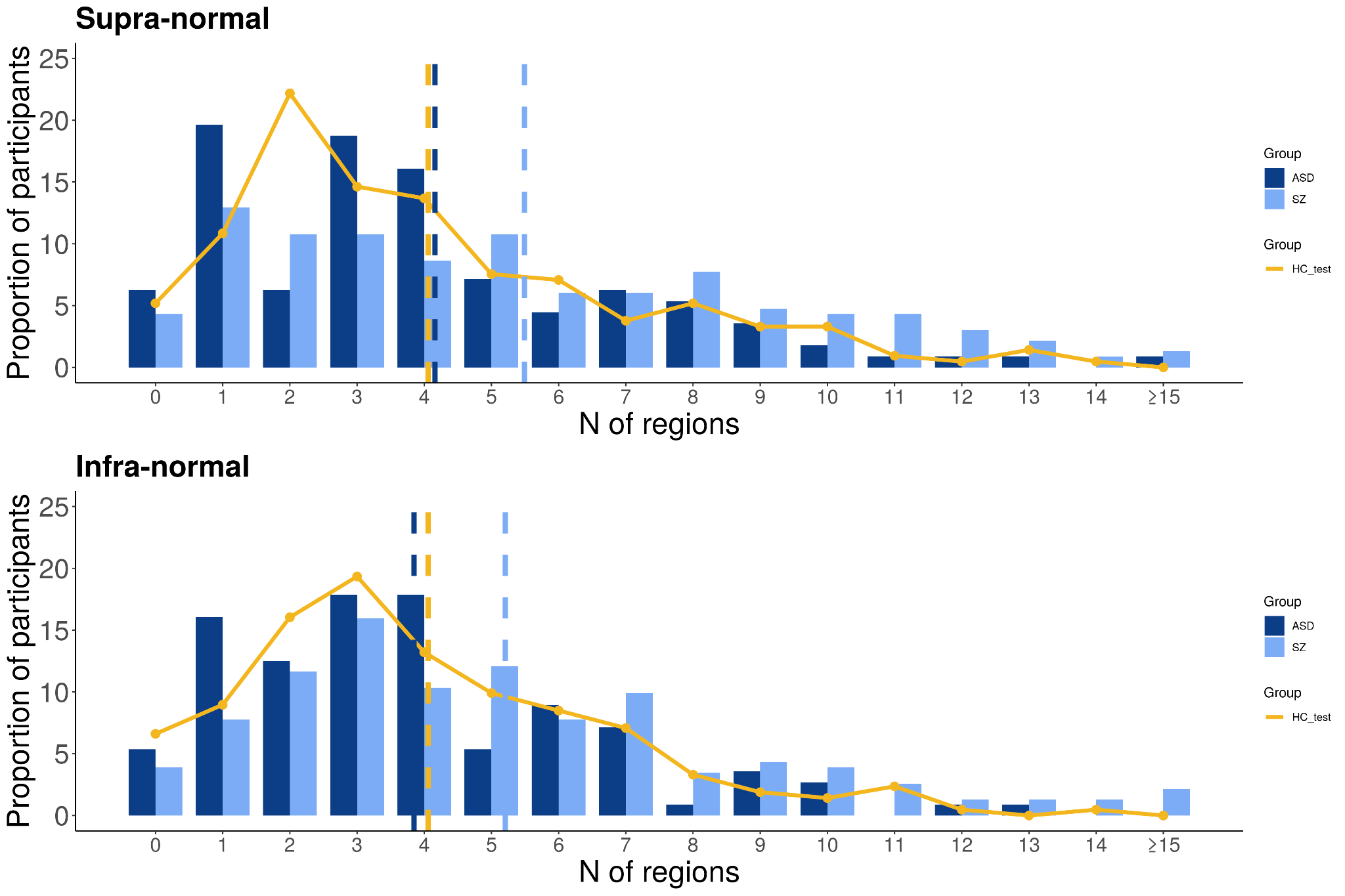

Supplemental Figure 19. Distribution of the number of regions with infra- or supra-normal deviance per male participant. Bar plots and curves display the distribution of the proportion of male individuals per amount of regions with supra-normal and infra-normal deviations. HC_test_, healthy individuals from the test set; ASD, Autism Spectrum Disorders; SZ, schizophrenia.

*Multivariate analysis*

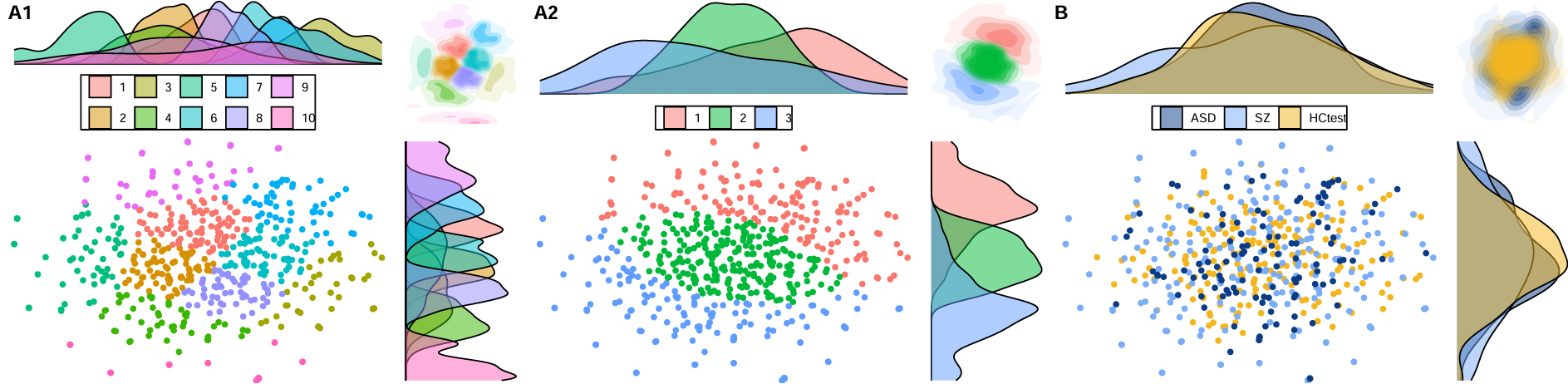

Supplemental Figure 20. Panels A1 and A2 present the optimum (A1) and three cluster results (A2) of k-medoid clustering applied to the 2D embedding of z-scores for regional cortical thickness AI, generated using tSNE. Panel B demonstrates that these clusters did not yield any meaningful differentiation based on diagnosis. AI, Asymmetry Index; HC_test_, healthy individuals from the test set; ASD, Autism Spectrum Disorders; SZ, schizophrenia.

**References**

1) [23774715](http://www.ncbi.nlm.nih.gov/pubmed/23774715)

2) [28291247](http://www.ncbi.nlm.nih.gov/pubmed/28291247)

3) 23479657

4) 32015023

5) 30569528

6) 27550776

7) 24736181

8) 32964935

9) 36920290

10) 33741990

11) 25412575

12) 21051551

13) <http://brain-development.org/ixidataset/>

14) 34584100

15) 30015807

16) 19929323

17) 23087608
